# Proteomics Reveal Clusters of Hypertension Cases Associated with Differing Prevalence of Cardiovascular and Renal Complications

**DOI:** 10.64898/2026.03.03.26347534

**Authors:** Yani Pehova, Simone Apella, Dmitry Kolobkov, Andrzej R. Malinowski, Marcin Pawlowski, Mark A. Strivens, Jason Sardell, Steve Gardner

## Abstract

**Background:** Hypertension affects over 30% of adults and is the leading risk factor for cardiovascular disease. It often presents without obvious symptoms, meaning that, although effective therapies exist, hypertension remains widely undiagnosed and insufficiently treated. Genomics-based prediction methods have shown only modest benefits for these disorders, but proteomic markers have demonstrated potential for greater predictive and clinical value.

**Methods:** We applied a novel machine-learning based patient stratification analysis pipeline to proteomics data for 7,086 hypertension patients from UK Biobank’s Pharma Proteomics Project cohort (2,911 proteins). We evaluated the contribution of each protein to the output of a tree-based risk model to explore the combinations of protein expression values that naturally separate hypertension cases into clusters and assessed the prevalence of cardiovascular and renal complications within each obtained cluster.

**Results:** We identified 10 clusters of hypertension patients segregated by differential expression of HAVCR1, PLAT, PTPRB, REN and RTN4R. Four of these clusters showed statistically significant enrichment for cardiovascular and renal complications, and three of them had significantly lower prevalence of complications than expected among hypertension patients.

**Conclusion:** We hypothesize that the hypertension clusters identified may represent distinct mechanistic subtypes. With further study this could help focus studies on subgroups of hypertension patients with a shared disease etiology, identify more personalized precision medicine treatment options for each subgroup, and develop mechanism-based biomarker tests to support enriched clinical trial recruitment.

## 1 Introduction

Hypertension is the most common risk factor for cardiovascular disease and one of the leading causes of premature mortality worldwide. Recent studies estimate that 31% of adults, approximately 1.8 billion people globally, have hypertension [1]. A significant proportion of them are unaware of their condition due to the “silent” nature of the disease with symptoms that are not normally noticeable [2].

Despite the availability of effective treatments, hypertension therefore remains underdiagnosed and undertreated, resulting in greater risk of developing life-threatening late-stage cardiovascular and renal complications. Later diagnosis and initiation of treatment results in progressive organ damage and more serious forms of disease that are more expensive to treat.

Hypertension has diverse clinical presentations, multiple underlying disease mechanisms, and patients exhibit variable responses to treatment [3, 4]. This is, in part, due to the heterogeneous nature of the condition. A greater understanding of the disease mechanisms underpinning hypertension, related cardiovascular diseases, and an individual’s risk of developing these cardiovascular and renal complications is essential for developing more effective, personalized approaches to disease management and clinical treatment.

Many clinical risk scores have been developed to predict the risk of cardiovascular disease [5–7]. One example is the Framingham Hypertension Risk Score that assesses the 4-year risk of developing hypertension in normotensive individuals [7]. These traditional risk scores rely on regression-based modelling of clinical and demographic features, including age, sex, cholesterol levels, blood pressure, presence of comorbidities such as diabetes, family history, and smoking status.

Although the current gold standard, these scores are often not directly transferable to diverse cohorts, requiring recalibration [8–10] and they may fail to capture the underlying disease biology or the dynamic changes to an individual’s health and environment. Such scores are potentially useful for assessing a person’s relative risk of hypertension within a population health setting, but they do not allow clinicians to predict the types of complications an individual patient is likely to experience or to stratify patients by mechanistic or complication subtype to help select the most appropriate treatments for individual patients.

### 1.1 Genomics vs Proteomics

Genome-wide association studies (GWAS) of multi-ancestry cohorts have identified more than 1,000 loci associated with various blood pressure traits [11, 12]. Polygenic risk scores (PRS) derived from these GWAS studies aim to provide individualized risk scores and have been shown to moderately improve risk prediction when integrated with clinical and demographic features [13–15]. However, because hypertension and cardiovascular diseases are complex, involving multiple alleles and genes interacting with each other and multiple exogenous factors, PRS are poorly suited to evaluating personal risk and making personalized recommendations based on the underlying mechanistic drivers of disease, especially across different ancestries [16].

PRS use the individual’s genome, which is a static measure, and builds scores from millions of infinitesimal, linearly additive, single SNP contributions, which do not change over a patient’s lifetime. They cannot reflect the full multifactorial nature of hypertension and its complications or identify the specific mechanisms most responsible for an individual patient’s disease risk. They can also struggle to be portable across populations, especially of different ancestry [16, 17].

Recent advancements in high-throughput proteomics have enabled the collection of large-scale plasma proteomes, offering an opportunity to gain new molecular insights into hypertension and its consequences. Plasma proteomics can provide insight into an individual’s current health status at the time of sampling, reflecting the combined impacts of genetics, comorbidities, lifestyle and environment, and the effects of drug therapies, thereby potentially enhancing patient stratification and precision medicine.

Phenotype-predictive models trained on UK Biobank (UKB) Pharma Proteomics Project (UKB-PPP) plasma proteomics dataset [18] plus baseline covariates substantially outperformed PRS on average across multiple phenotypes [19, 20]. Other studies have used the same UKB-PPP dataset to predict disease risk while simultaneously uncovering the biological pathways and individual protein-to-disease links that drive model performance [20, 21].

A Qatari Biobank study collected measures of 1,300 plasma proteins for 778 individuals and applied regression-based modelling to identify 13 proteins associated with hypertension and 25 proteins associated with mean arterial pressure [22]. A study on the German KORA cohort of approximately 1,500 individuals identified 49 proteins that are associated with hypertension [23]. A recent study of genetically predicted proteomic signal in cardiovascular disease using proteome-wide Mendelian randomization supported by genetic colocalization analysis inferred ACOX1, FGF5, FURIN and MST1 to be causally linked with blood pressure, risk of stroke, and coronary artery disease (CAD) [24].

These studies looked for direct associations between the circulating proteins and the condition, assuming a binary case:control phenotype classification, and thus failing to account for any underlying disease subtypes in patient populations. This approach is unlikely to yield truly actionable results as it ignores the cohorts’ underlying heterogeneity. For example, subgroups of hypertension patients exhibit heterogeneous levels of renin [25], and proteomic studies have suggested conflicting associations between plasma renin and hypertension [26]. Such discrepancies may arise from a lack of awareness of the underlying disease subgroups, which may have different mechanistic etiologies that may affect their proteomic profiles.

This highlights the need for unbiased approaches that better account for the structure of patient populations. Combining studies of the proteomically distinct subgroups of hypertension patients with in-depth knowledge of the effect of therapies on protein expression such as [27] can lead to the development of highly effective personalized treatments.

Hypertension subtypes have been previously identified using clinical and demographic features, with isolated diastolic, isolated systolic, and systolic-diastolic hypertension being three commonly considered subtypes [3, 28]. A genomic study on FinnGen identified four genetic hypertension components (obesity, dyslipidemia, hypolipidemia, and short stature) based on just 73 variants which the authors suggest could serve as the basis for stratification [29]. Another study integrating genomic and transcriptomic data identified two subgroups of hypertension patients based on whether hypertension-associated molecular alterations are diffused or localized [30]. None of these has yet generated clinically actionable results.

Multi-omics data, including metabolomics, were recently used in machine learning models to predict one of four pre-defined hypertension subtypes – primary hypertension, endocrine hypertension due to primary aldosteronism, pheochromocytoma/functional paraganglioma, or Cushing syndrome – in the presence of controls [31]. Some proteomics-based stratification studies of other phenotypes are present in the literature showing subtypes of acute myeloid leukemia [32] and soft tissue sarcoma [33]. Proteomic data of 470 patients at Imperial College NHS Trust with pulmonary hypertension was analyzed [34], identifying four clusters with diverse survival rates [34].

In this study we used the large UKB-PPP cohort [18], comprised of approximately 53,000 individuals to reveal diverse and biologically substantiated hypertension subtypes associated with differential complication risks [18]. We developed and applied a novel approach to patient stratification, which considers a machine learning risk score model and uses its structure to infer phenotype subtypes by leveraging unsupervised clustering techniques. To the best of our knowledge this paper presents the first large-scale proteomics-based stratification study of hypertension and its potential complications.

## 2 Methods

We analyzed Olink proteomics data collected at initial assessment visit from 53,073 individuals from the UKB-PPP [18], covering 2,923 plasma proteins at the time of analysis (October 2023). We first removed 0.5% proteins with the highest missingness ratio (greater than 20%), and then we removed 0.5% participants with the highest missingness ratio (greater than 63%) leaving 2,911 proteins and 52,806 participants. The proteins removed are listed in Supplementary Data (Table S5). Additionally, 27 UKB participants who withdrew consent after initial QC were removed from the dataset in August 2025, leaving 52,773 participants.

### 2.1 Study Design

We identified 17,952 hypertension cases with ICD-10 code I10 (Essential (Primary) Hypertension). This cohort of cases was split according to the time of I10 diagnosis and only the 11,252 cases where the I10 diagnosis predates the date of initial assessment visit were kept in order to isolate cases with existing hypertension at the time of proteomics sample. Cases who indicated that they were taking angiotensin-converting enzyme (ACE) inhibitors at the time of their initial assessment visit were removed as the medication is known to affect the expression of two key proteins that were assigned high importance in initial analyses: ACE and renin (REN) [35]. After this, 7,086 cases remained.

Controls were selected with a target case:control ratio of 1:2. To ensure properly characterized control subjects, samples with any of the ICD-10 codes I10-I15 were excluded. Participants receiving blood pressure medication and participants exhibiting systolic blood pressure outside of the range 90-135 mmHg or diastolic blood pressure outside of the range 60-85 mmHg at initial assessment visit were also excluded. The UK National Health Service (NHS) standard for normal blood pressure, which is between 90/60 mmHg and 120/80 mmHg [36], was originally considered, but due to a shortage of controls this was relaxed to between 90/60 mmHg and 135/85 mmHg. A full description of the UKB data fields and codings used can be found in Supplementary Data, Table S1.

Sex- and age-matching was performed when subsampling controls to reach the target ratio. Only 16,047 hypertension controls were available for this study, 13,016 of which had to be included in the dataset to obtain the desired 1:2 case:control ratio. This limited our ability to select sex- and age-matched controls, resulting in slight differences in the distribution of age and sex among cases and controls (see 2.2.3 Model Training and Supplementary Data, Table S3).

The genetic ancestry of samples was computed using GRAF-pop [37] revealing that 86.1% of samples have European ancestry. Of all participants, 86.6% self-identified to be of White British ethnic background. For a detailed ancestry and ethnicity breakdown see Supplementary Data, Figure S1.

Indications representing complications of hypertension were selected based on established clinical guidelines and literature evidence [38, 39]. These included atrial fibrillation, myocardial infarction (MI), chronic ischemic heart disease (CIHD), heart failure, stroke, cardiac arrest, cardiomyopathy, and renal complications. Diagnostic codes corresponding to each condition were identified using ICD-10 classifications and detailed definitions and codes, together with a full summary of our study design, are included in the Supplementary Data, Table S1.

### 2.2 Analysis

We employed a machine-learning approach to construct a patient stratification pipeline based on the Olink proteomics data. For our hypertension dataset, we trained an XGBoost classifier [40] predicting the case/control status of input samples from proteomics data (the compute environment specifications can be found in Supplementary Data, Model Training, System Specifications).

XGBoost was selected due to its capability to capture non-linear interactions between input features, and its inherent ability as a decision tree model to separate its input space by identifying meaningful thresholds in the input features and naturally separating groups of input samples that are “similar”.

Since the blood plasma proteome is affected by a multitude of factors, most of which are unrelated to a single phenotype [41], clustering patients based directly on normalized protein expression may yield clusters of little relevance to the phenotype. For this reason, we considered the space of per-sample contributions of each protein to the output of the trained XGBoost classifier, in which a greater portion of the variance is expected to correspond to the phenotype. An example showing the relationship between HAVCR1 and hypertension and the differences between expression and model contribution is shown in Figure 1.

**Figure 1.**
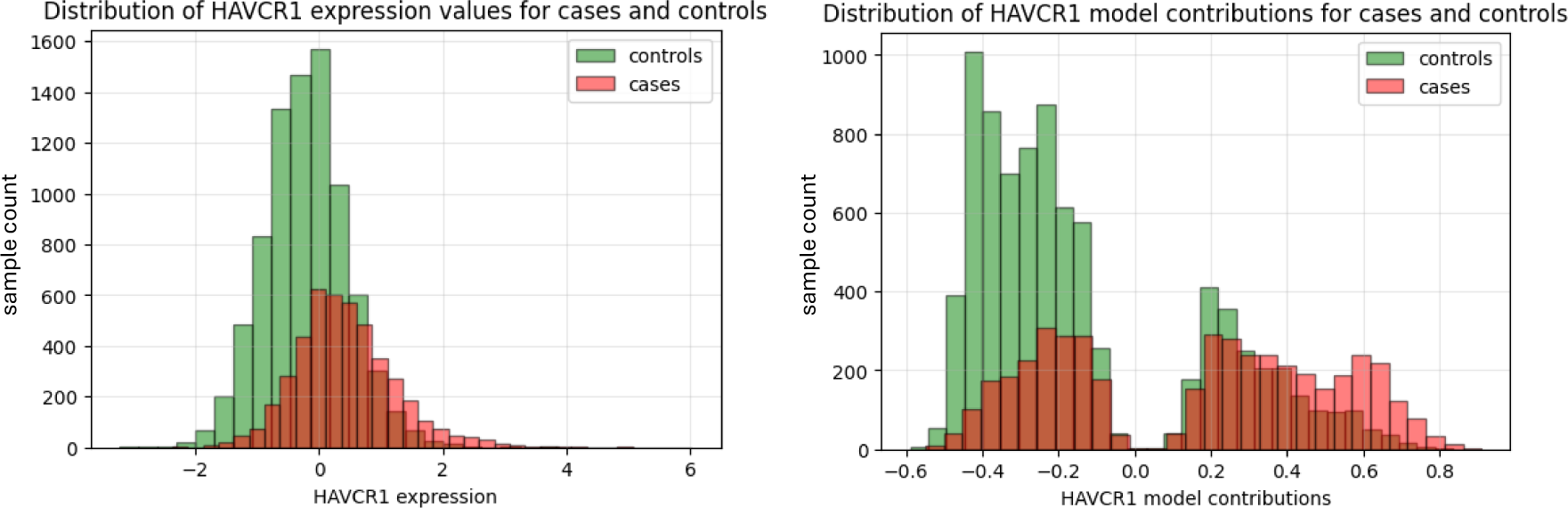
Model contribution values create more explicit case-control and patient subgroup separation compared to normalized expression values of HAVCR1 in our hypertension dataset. In the left histogram, mean expression of HAVCR1 is higher in cases than controls, but the distributions overlap considerably and exhibit no clear separation between groups of cases. In contrast in the histogram on the right, two distinct clusters of hypertension cases are represented as distinct peaks based on relative contributions of HAVCR1 to the model output.

Further to stratifying our cohort of hypertension cases, we explored connections between the expression of a handful of key “segregating proteins” used for the stratification and the severity of the phenotype.

#### 2.2.1 Data Processing

Quality controlled proteomics data, age, and sex were loaded as input features for the samples from our hypertension dataset and standardized. Inputs were not imputed as XGBoost models allow for missing values.

#### 2.2.2 Hyperparameter Tuning

Before training our XGBoost models used to generate the stratification results in this manuscript, a 3-fold Bayesian hyperparameter search [42] with 20 iterations was performed to choose optimal training parameters (see Table 1). The algorithm explores multiple combinations of XGBoost parameters including learning rate, maximum tree depth, regularization strength and number of estimators. Each parameter combination was evaluated, identifying the parameters that maximized the average test AUROC score across the three folds (also referred to as validation score). Details of the hyperparameter search space are described in Supplementary Data, Model Training, XGBoost Hyperparameter Search Space.

**Table 1.** Performance metrics of XGBoost models predicting hypertension case/control status from proteomics data across five folds. The optimal hyperparameters identified with hyperparameter search were: n_estimators=2000, max_depth=3, learning_rate=0.0932, reg_alpha=12.7671.

| Metric | Value |
| --- | --- |
| Train AUROC | 1.00 ± 0.0000 |
| Test AUROC | 0.89 ± 0.0026 |
| Train AUROC with age, sex | 1.00 ± 0.0000 |
| Test AUROC with age, sex | 0.89 ± 0.0029 |

To ensure comparability of train and test metrics in the presence of covariates, the same set of hyperparameters was used both in runs with and without age and sex as covariates, and across folds during cross-validation. To make this possible, the hyperparameter search was performed only once, on the Train dataset (for definitions of our Train and Test datasets see 2.3.1 Data Splitting) without covariates, as opposed to the training data of each fold, or separately for data with and without covariates. This is a standard approach as hyperparameter searches usually employ a *k*-fold procedure while ultimately retraining a final model on the entire dataset. More details on the training routine and data splitting employed during this step are described in 2.3.1 Data Splitting below.

#### 2.2.3 Model Training

The optimal parameters identified in the previous step were used to train XGBoost classifiers in a five-fold cross-validation loop (described in more detail in 2.3.1 Data Splitting below) predicting the case/control status of input samples from proteomics data.

As blood plasma proteomics may be confounded by many factors, a major focus of the methodological strategy of this paper was controlling for age and sex as covariates, both of which are known to strongly affect protein expression [43–46]. This was necessary to increase the likelihood that clusters reflect real differences in disease biology that can potentially be used to identify precision medicine treatments and not simply clustering of hypertension cases by age and sex, or other covariates.

Cases and controls were age- and sex-matched to the extent possible when generating our hypertension dataset, so age and sex features are not generally expected to be used by our models unless their joint distributions with particular proteins and the phenotype are skewed. However, due to a shortage of controls, some age and sex differences remained (see Supplementary Data, Table S3). We therefore further controlled for covariates by including them as input features in the model.

Metrics such as train and test AUROC score were recorded to evaluate the performance of the trained models. A benchmark model without covariates was trained with the same hyperparameters to showcase the contribution of covariates to the reported model accuracy metrics.

#### 2.2.4 Methods for Stratification

Following model training, the contribution of each input protein to the model prediction for each sample was collected in an array, restricting the dataset only to cases according to the supplied case/control labels (rather than the model predictions). Each sample in this array was mapped to 2 dimensions using Uniform Manifold Approximation and Projection (UMAP) [47], after which multiple iterations of the spectral clustering algorithm [48, 49] with default parameters, 10 *k*-means initializations and Radial Basis Function (RBF) kernel with coefficient 1 were applied to select a set of “segregating proteins” for the final patient stratification.

After the segregating proteins were identified, a final round of spectral clustering was performed on these segregating proteins only, again with default parameters, to produce the final stratification of the model’s training data. The number of clusters was chosen manually based on their shape and appearance and to ensure a minimum cluster size of approximately 100 samples, and the silhouette score [50] was reported as a quality metric.

The silhouette score measures how well each patient fits within their assigned subgroup by comparing the average distance to patients in the same cluster against the average distance to patients in the nearest distinct cluster. This score ranges from −1 to 1, where values close to 1 are indicative of well separated and distinct clusters, and values close to 0 or negative suggest that clusters are not well separated and may even overlap.

To produce a map that can be applied to stratify external data, the trained model, protein contribution calculation, and UMAP dimensionality reduction described above were composed into a single map. Due to the spectral clustering algorithm’s reliance on input data, to produce the final step, a *k*-nearest neighbor classifier with *k*=10 was fitted to the training data’s 2-dimensional coordinates after dimensionality reduction and corresponding cluster assignment. For each of the five folds, the resulting end-to-end stratification map was applied to the fold’s test data to produce a stratification of patients not seen during training. The same end-to-end stratification map was further applied to our hold-out Test dataset for the purposes of assessing stability, as described in 2.3.3 Stratification Stability.

#### 2.2.5 Enrichment Analysis for Common Hypertension Complications

After obtaining the stratification of hypertension cases, we analyzed the differential expression of segregating proteins across all clusters and explored their clinical significance by connecting them to higher or lower prevalence of common hypertension complications.

First, we evaluated the proteomic profile of each cluster by applying a Mann-Whitney U test [51] to the differential expression of each protein within the cluster versus its expression in all cases and all controls. For each of the comparisons, we reported Cohen’s d effect size, which measures the standardized mean difference between the two compared distributions. Cohen provided the conventions for defining small (0.2), medium (0.5), and large (0.8) effect sizes for computing statistical power [52]. In the context of protein expression analysis, a positive Cohen’s d indicates that a protein is up-regulated in the cluster of interest, whereas a negative Cohen’s d indicates down-regulation.

We evaluated which complications are significantly enriched within each cluster using one-tailed Fisher’s exact tests [10]. Clusters with fewer than 100 samples were skipped during this analysis due to the small sample size and expected even smaller sample size of the cluster in test data. A cluster was classified as enriched for a complication if the odds ratio of the complication for the cluster relative to the remaining hypertension cases was greater than 1.5 and the enrichment was significant at the *p* < 0.05 level after applying Benjamini-Hochberg (BH) false discovery rate (FDR) correction for the total number of tests across all complications and clusters.

The cutoff of 1.5 was chosen to represent strong enrichment. A cluster was classified as “protected” against a complication if it had an odds ratio less than 0.67 and *p* < 0.05 after applying BH correction. The cutoff of 0.67 was chosen as the inverse of 1.5 to represent the analogous strength of enrichment in the opposite direction.

We evaluated whether clusters that were enriched for or protected against at least one complication also differed significantly in sex ratio or age from other clusters using Fisher’s exact test for sex and a t-test for age. For sex, odds ratios greater than 1 indicate male-enriched clusters and odds ratios smaller than 1 indicate female-enriched clusters. Age- and sex-enrichments were reported if they were significant at the *p* < 0.05 level after applying BH correction over all clusters, and we further filtered the results for sex to report only associations with odds ratios smaller than 0.67 or greater than 1.5.

To further validate the identified cluster enrichments in the presence of age and sex, for each patient cluster and each complication, we fitted a logistic regression model using cluster membership to predict complication status with age and sex as covariates to remove any residual covariate bias. As a measure of the age- and sex-agnostic enrichment of the cluster for the complication, we reported the Wald *p*-value of the cluster membership variable. This multi-stage enrichment validation procedure ensures our analysis is minimally confounded by age and sex in the presence of a limited pool of controls.

The same differential expression and enrichment analysis was repeated for the stratification of each fold’s test data obtained as described in 2.2.4 Methods for Stratification to assess how the results generalize to external data and to what extent they may be artifacts of the model’s training data. Unlike for training data, clusters with fewer than 100 samples were not skipped during this analysis as enrichments using test data serve as follow-up validation of enrichments using train data.

### 2.3 Replication and Validation

Throughout our analysis, the prediction accuracy and stability of our models was continuously validated using UKB samples not involved in model training.

#### 2.3.1 Data Splitting

The input data *X* and phenotype vector *y* for our hypertension dataset – consisting of 20,102 samples and 2,913 features (2,911 proteins plus age and sex) – were split into datasets Train = (*X_train_*, *y_train_*) and Test = (*X_test_*, *y_test_*) in a 4:1 ratio, stratified by *y*. The number of samples in each portion of the dataset are shown in Supplementary Data, Table S4. A 3-fold Bayesian hyperparameter search with 20 iterations was then performed on *X_train_*, *y_train_* to select the set of XGBoost hyperparameters that yielded the best validation AUROC score (details on the search space are described in Supplementary Data, XGBoost Hyperparameter Search Space).

A five-fold cross-validation training loop was then performed on *X_train_*, *y_train_*, assessing the train and test AUROC for each of the resulting five models. Each model yielded a stratification of its train data and a corresponding stratification map assigning a cluster label which may be applied to external data. A summary of the data splitting procedure and how it fits within the steps of our analysis can be found in Figure 2a and b.

**Figure 2.**
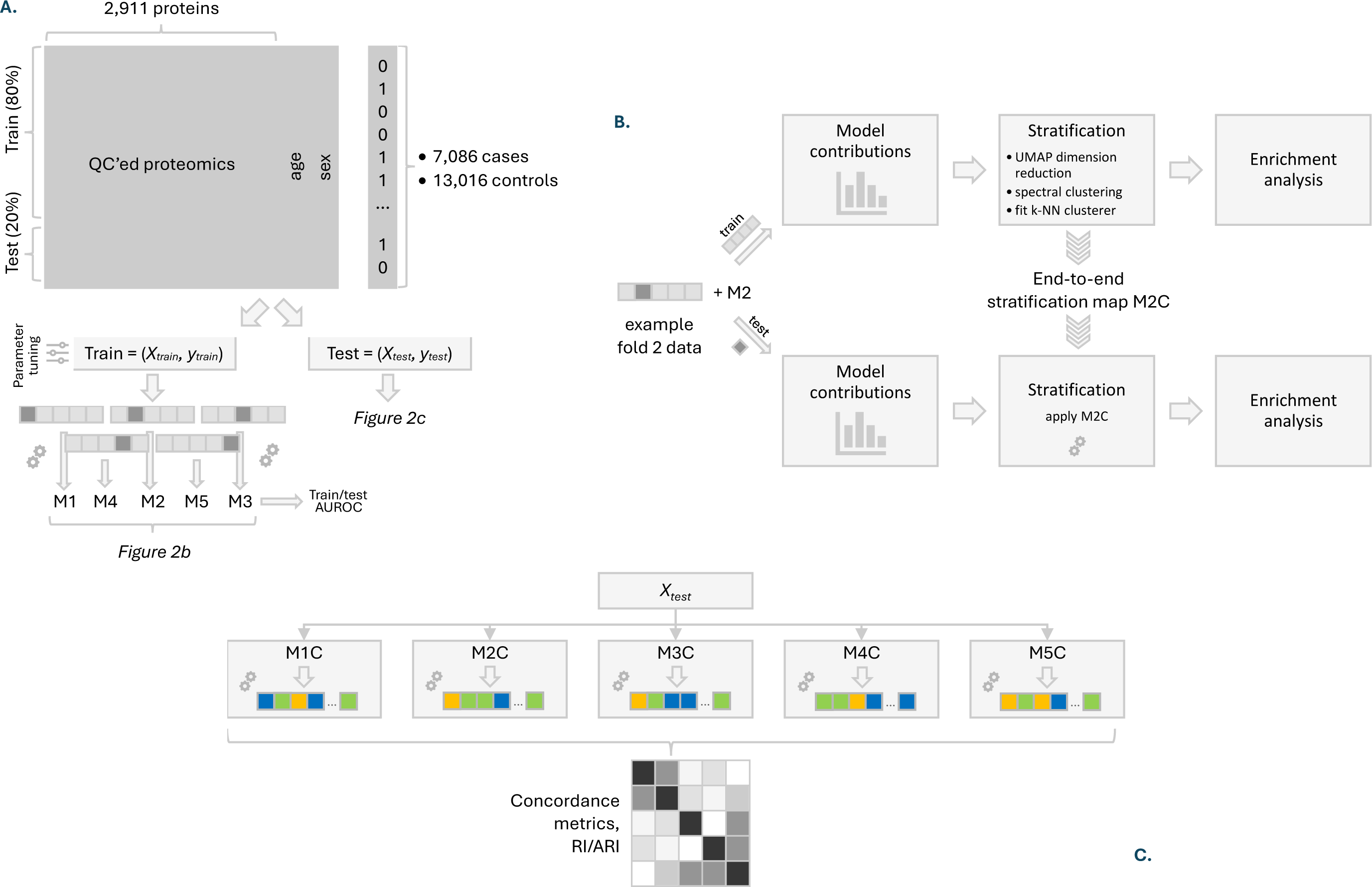
**A.** Schematic of data sources, data splitting and subsequent model training in a five-fold cross-validation loop. Shaded sections depicted within each fold denote the fold’s test data. The resulting five models trained on each of the five folds have been abbreviated as M1-5. **B.** Analysis pipeline performed on the model resulting from each fold in the five-fold cross validation loop. Train data from the fold yields an end-to-end stratification map, which can then be applied to test data. Pictured is fold 2, trained model M2, and the resulting stratification map M2C. **C.** Stability analysis assessing the pairwise concordance of cluster labels (illustrated by different colors) of samples in our hold-out test dataset *Xtest* produced by each of the five stratification maps M1-5C.

#### 2.3.2 Stratification

For each of the five folds, the resulting stratification map was applied to the fold’s test data to produce a stratification of hypertension cases not seen during training. Differential expression and enrichments for complications, age, and sex were recalculated on this test data to assess how they generalize to external data and to what extent they may be artifacts of the model’s training data. Enrichments for complications, age, and sex were considered significant if they had a BH-corrected *p*-value < 0.05.

#### 2.3.3 Stratification Stability

To assess stability, the stratification maps from each of the five folds were applied to the cases in our Test dataset, yielding five stratifications (also referred to as labellings) of *X_test_*. To evaluate stratification reproducibility and its sensitivity to the exact training data included in different folds, the Rand Index (RI) [11] and Adjusted Rand Index (ARI) [12, 13] were computed for all ten pairs of labellings to measure their concordance.

The RI of two labellings measures the proportion of sample pairs for which both labellings agree on whether the samples belong to the same cluster or different clusters. The RI is between 0 and 1 and is 1 if and only if the stratification assignments are identical. The ARI, valued between −0.5 and 1, includes a correction for expected random agreement. For comparison, a random binary classification model would achieve an AUROC score of 0.5, so if AUROC were to be normalized in the same way, an AUROC score of 0.5 would map to 0 and an AUROC score of 1 would map to 1. A schematic of our stability analysis can be found in Figure 2c.

## 3 Results

### 3.1 Model Performance

We trained XGBoost models in a five-fold cross-validation loop, predicting the case/control status of samples as a function of input protein expression data with age and sex as covariates. The parameters used by each model and resulting mean and sample standard deviations of train and test AUROC scores are summarized in Table 1. Further model metrics are listed in Supplementary Data, Table S6. We observed an extremely high diagnostic AUROC score of 0.89 with standard deviation across folds of 0.0026. Performance was not improved by including age and sex as covariates in the model.

### 3.2 Stratification

Each trained model underwent a feature selection step selecting a set of significantly segregating proteins that were used for stratification. We present the sets of segregating proteins used in each fold within our cross-validation loop in Table 2, and a single example set of stratification results for one of the five models trained (fold 1), while the remaining stratification results can be found in Supplementary Data, Additional Stratification Results. The proteins HAVCR1 and REN appeared consistently as segregating proteins in all five folds (shown in bold in Table 2) while others such as PON3, NTproBNP, GDF15 and PTPRB appeared either 2 or 3 times.

**Table 2.** Segregating proteins obtained by applying our patient stratification pipeline to the sets of hypertension cases in each of the five cross-validation folds. Segregating proteins are subsequently used to identify subgroups of hypertension cases. The proteins shown in bold are segregating for all five folds.

| Fold 1 | Fold 2 | Fold 3 | Fold 4 | Fold 5 |
| --- | --- | --- | --- | --- |
| **HAVCR1** | GDF15 | **HAVCR1** | CHI3L1 | ACTA2 |
| PLAT | **HAVCR1** | NPL | GDF15 | **HAVCR1** |
| PTPRB | **REN** | NTproBNP | **HAVCR1** | NTproBNP |
| **REN** |  | PON3 | PON3 | PON3 |
| RTN4R |  | PRSS8 | **REN** | PTPRB |
|  |  | **REN** |  | **REN** |

The cohort of training hypertension cases from fold 1 yielded 10 clusters defined by the five segregating proteins HAVCR1, PLAT, PTPRB, REN and RTN4R with silhouette score of 0.59, denoting good clustering quality (see Figure 5). The differential expression of each cluster was evaluated against cases and controls (see Figure 3). The Cohen’s d effect sizes for each cluster and protein are shown in Figure 4 and in more detail in Supplementary Data (Table S7). We further analyzed each cluster to evaluate the prevalence of common complications of hypertension and its age and sex distribution (see also Figure 5). We observed higher prevalence of cardiovascular and renal complications in clusters 3, 4, 8 and 10 and lower prevalence in clusters 2, 6 and 9 compared to the underlying prevalence among all hypertension cases.

**Figure 3.**
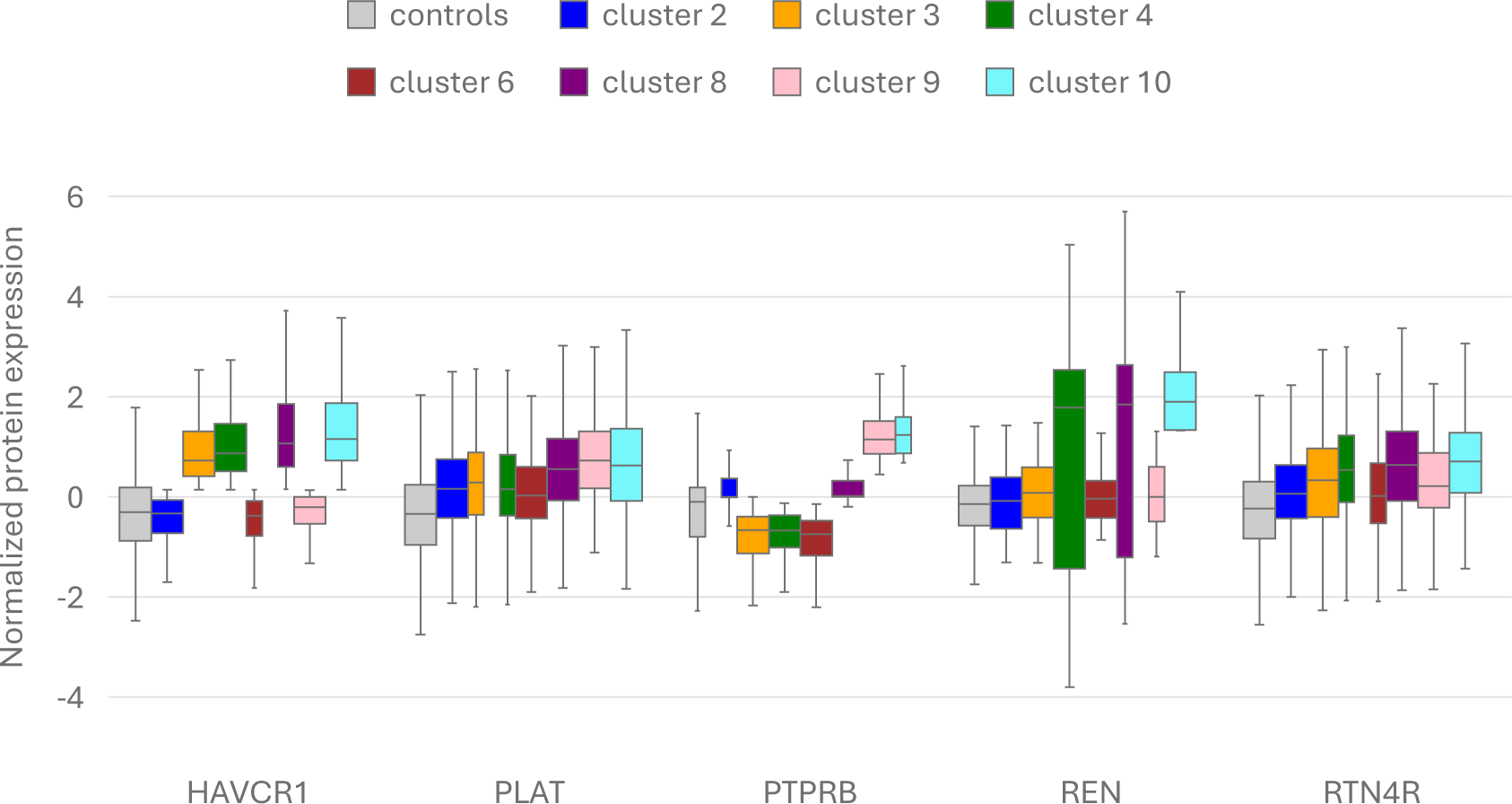
Differential expression of each of the five segregating proteins for fold 1 among clusters of hypertension cases from the fold’s training data. Pictured are the middle three quartiles. Each whisker extends to the farthest data point lying within 1.5x the inter-quartile range (IQR) from the corresponding box. Remaining folds and results on test data can be found in Supplementary Data, Additional Stratification Results.

**Figure 4.**
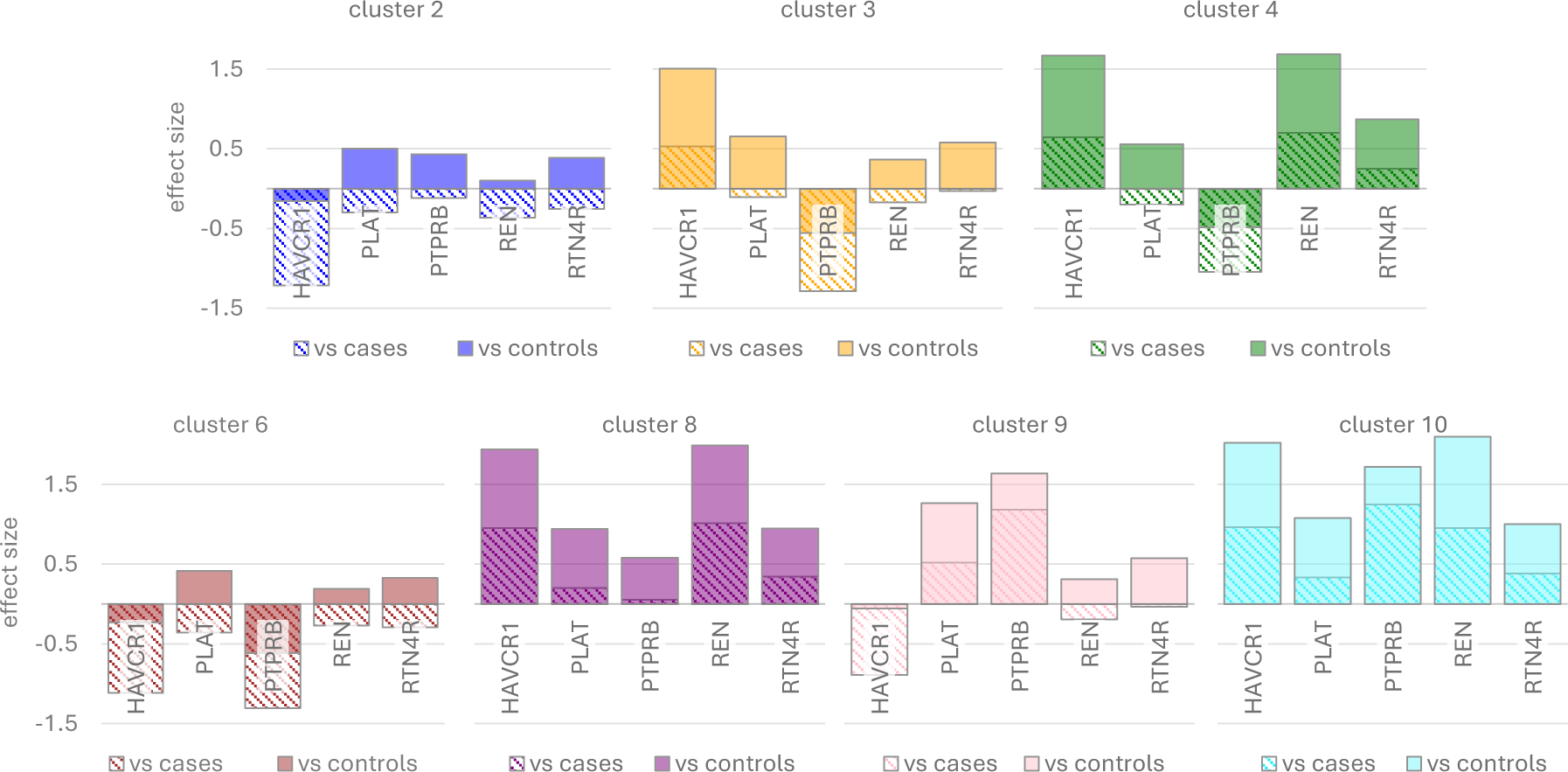
Cohen’s d effect sizes of segregating protein expression of hypertension cases in each cluster versus the cohort of all cases or controls, calculated using the segregating proteins and cluster labels given by fold 1, and the training data from fold 1. Remaining folds and results on test data can be found in table format in Supplementary Data, Additional Stratification Results.

**Figure 5.**
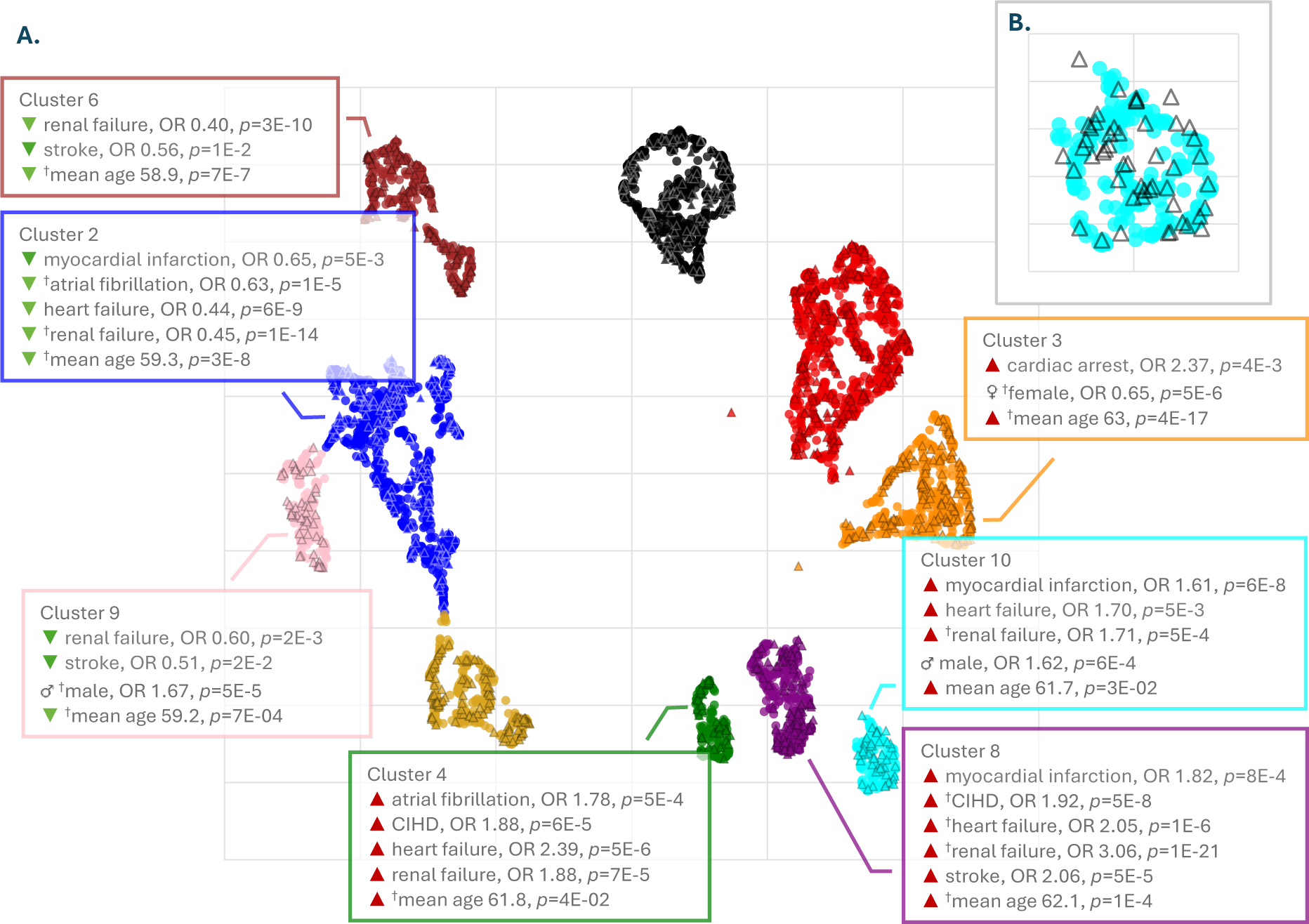
**A.** Cluster enrichments for complications, age and sex among hypertension cases from fold 1. Training data is shown as circle markers and test data is shown as triangles. Enrichments marked with a dagger (†) persisted among test data meaning that they were significant at the *p* < 0.05 level after applying Benjamini-Hochberg FDR correction. Remaining folds can be seen in Supplementary Data, Additional Stratification Results. **B.** A closer look at the overlay of test data (triangles) on train data (teal circles) for cluster 10 is shown in the top right-hand corner.

Clusters 4, 8 and 10 were most enriched for complications and showed significantly elevated expression of REN and HAVCR1compared to controls (effect sizes > 1.6, *p* < 10^-16^). PTPRB and PLAT were up-regulated in all three high-risk clusters but with different effect sizes. Cluster 4 showed a slight up-regulation of PLAT (effect size 0.56, *p* = 10^-13^) and weakly down-regulated PTPRB levels (effect size −0.48, *p* = 2 x 10^-15^) compared to controls. Cluster 8 exhibited up-regulated levels of PLAT (effect size 0.94, *p* = 3 x 10^-59^), and weakly up-regulated levels of PTPRB (effect size 0.58, *p* = 2 x 10^-29^). Cluster 10 also showed PLAT up-regulated levels (effect size 1.08, *p* = 4 x 10^-39^), but strongly up-regulated PTPRB levels (effect size 1.72, *p* = 10^-100^).

Clusters 2, 6 and 9 were at low overall risk of complications and showed expression of REN and HAVCR1 close to controls (effect sizes smaller than 0.35 in absolute value and *p*-values of the order 10^-6^ or greater). All three low-risk clusters showed higher levels of PLAT and RTN4R compared to controls, but with different effect sizes. PTPRB expression compared to controls differs among the low-risk clusters. Cluster 2 exhibited weak up-regulation of PLAT (effect size 0.5, *p* = 2 x 10^-47^), and up-regulation of RTN4R (effect size 0.39, *p* = 3 x 10^-27^). PTPRB was weakly up-regulated in cluster 2 (effect size 0.43, *p* = 4 x 10^-37^). Cluster 6 exhibited weak up-regulation of PLAT (effect size 0.42, *p* = 6 x 10^-17^), and up-regulation of RTN4R (effect size 0.32, *p* = 7 x 10^-11^). Contrary to cluster 2, cluster 6 showed a weak down-regulation of PTPRB (effect size −0.62, *p* < 3 x 10^-41^). Cluster 9 showed up-regulation of PLAT with higher effect size compared to clusters 2 and 6 (effect size 1.27, *p* = 2 x 10^-71^), and higher levels of RTN4R (effect size 0.58, *p* = 10^-18^). Contrary to clusters 2 and 6, cluster 9 showed strong up-regulation of PTPRB (effect size 1.63, *p* = 3 x 10^-123^).

Cluster 4 was significantly enriched for atrial fibrillation, CIHD, heart failure, and renal failure (OR = 1.78, *p* = 5 x 10^-4^), and cluster 8 was enriched for MI, CIHD, stroke, heart failure, and renal failure with extremely strong prevalence of renal failure at OR = 3.06, *p* = 10^-21^ and CIHD at OR = 1.92, *p* = 5 x 10^-8^. Cluster 10 was very strongly enriched for MI (OR = 1.61, *p* = 6 x 10^-8^) and also heart failure, and renal failure (both have OR > 1.7 and *p* < 5 x 10^-3^). Cases in cluster 3 were at risk of a narrower set of complications with only cardiac arrest being significant (OR = 2.37, *p* = 4 x 10^-3^).

Clusters 2, 6 and 9 appeared to be “protected” from complications (denoting negative correlation but not necessarily causality). Cluster 2 showed a lower prevalence of MI (OR = 0.65, *p* = 5 x 10^-3^), atrial fibrillation (OR = 0.63, *p* = 10^-5^), heart and renal failure (OR = 0.44, *p* = 6 x 10^-9^ and OR = 0.45, *p* = 10^-14^ respectively), while clusters 6 and 9 were both protected from renal failure and stroke. All reported enrichments are significant at the *p* < 0.05 level after applying a BH FDR correction.

Finally, each cluster enrichment was further validated in the presence of age and sex as covariates using a logistic regression model. The results are shown in Table 3. The majority of enrichments retained their significance when age and sex were factored out as covariates. Most significantly, both enrichments in cluster 9 and two out of three enrichments in cluster 10 did not remain statistically significant after controlling for age and sex. The corresponding analysis on test data is shown in Supplementary Data (Table S15).

**Table 3.** Wald test *p*-values and odds ratios for the logistic regression coefficient of each cluster in fold 1 as a predictor of each complication reported in Figure 5 on its own and in the presence of age and sex, among the fold’s training data. The *p*-values in the label, sex and age model shown in bold are significant at the *p* < 0.05 level after Benjamini-Hochberg FDR correction. Remaining folds and analogous results for test data can be found in Supplementary Data, Additional Stratification Results.

| Cluster | Complication | $p$-value<br>(label only) | Odds ratio<br>(label only) | $p$-value (label<br>+ sex + age) | Odds ratio (label<br>+ sex + age) |
| --- | --- | --- | --- | --- | --- |
| 2 | myocardial infarction | $9 \times 10^{-3}$ | 0.67 | $2 \times 10^{-2}$ | 0.70 |
| 2 | atrial fibrillation | $9 \times 10^{-6}$ | 0.63 | **$1 \times 10^{-3}$** | 0.71 |
| 2 | heart failure | $5 \times 10^{-8}$ | 0.45 | **$1 \times 10^{-6}$** | 0.49 |
| 2 | renal failure | $4 \times 10^{-14}$ | 0.45 | **$2 \times 10^{-11}$** | 0.49 |
| 3 | cardiac arrest | $2 \times 10^{-3}$ | 2.27 | **$5 \times 10^{-3}$** | 2.09 |
| 4 | atrial fibrillation | $5 \times 10^{-4}$ | 1.75 | **$2 \times 10^{-3}$** | 1.68 |
| 4 | CIHD | $4 \times 10^{-5}$ | 1.85 | **$5 \times 10^{-5}$** | 1.87 |
| 4 | heart failure | $1 \times 10^{-6}$ | 2.32 | **$4 \times 10^{-6}$** | 2.26 |
| 4 | renal failure | $5 \times 10^{-5}$ | 1.85 | **$2 \times 10^{-4}$** | 1.78 |
| 6 | renal failure | $1 \times 10^{-8}$ | 0.41 | **$5 \times 10^{-7}$** | 0.45 |
| 6 | stroke | $3 \times 10^{-2}$ | 0.58 | $6 \times 10^{-2}$ | 0.63 |
| 8 | myocardial infarction | $4 \times 10^{-4}$ | 1.78 | **$2 \times 10^{-3}$** | 1.66 |
| 8 | CIHD | $8 \times 10^{-9}$ | 1.89 | **$9 \times 10^{-7}$** | 1.74 |
| 8 | heart failure | $1 \times 10^{-7}$ | 2.02 | **$5 \times 10^{-6}$** | 1.86 |
| 8 | renal failure | $< 10^{-320}$ | 3.02 | **$< 10^{-320}$** | 2.85 |
| 8 | stroke | $1 \times 10^{-5}$ | 2.02 | **$7 \times 10^{-5}$** | 1.89 |
| 9 | renal failure | $5 \times 10^{-3}$ | 0.62 | $1 \times 10^{-2}$ | 0.65 |
| 9 | stroke | $5 \times 10^{-2}$ | 0.55 | $6 \times 10^{-2}$ | 0.56 |
| 10 | myocardial infarction | $5 \times 10^{-3}$ | 1.82 | $2 \times 10^{-2}$ | 1.67 |
| 10 | heart failure | $5 \times 10^{-3}$ | 1.67 | $2 \times 10^{-2}$ | 1.55 |
| 10 | renal failure | $4 \times 10^{-4}$ | 1.69 | **$2 \times 10^{-3}$** | 1.60 |

### 3.3 Replication and Validation

For each of the five folds, the stratification map returned by our pipeline was applied to the fold’s test data to produce a stratification of patients. The differential expression of the segregating proteins and all enrichments were validated on the test dataset. Test cases together with their clustering labels are denoted by triangular markers in Figure 5.

Differential expression of clusters from fold 1 on test data can be seen in Supplementary Data, Figure S2. Cluster enrichments of train data that replicated in test data are shown in Figure 5 (see Supplementary Data, Table S14 for *p*-values and odds ratios). In this context, the statistical significance of enrichments on test data is expected to be naturally lower due to the limited size of the test datasets. This is not necessarily a sign that enrichments don’t generalize but rather a natural consequence of the loss of statistical power when sample size is reduced. The corresponding results for the remaining four folds can be found in Supplementary Data, Additional Stratification Results.

We assessed the stability of our stratification pipeline by applying the five folds’ stratification maps to the hold-out Test dataset unseen by any of the models to measure their concordance. The unadjusted and adjusted Rand indices (RI and ARI) for each pair of models are shown in Table 4.

**Table 4.** Concordance of cluster labels obtained by applying each of the five cross-validation folds’ end-to-end stratification maps on our Test cohort of hypertension cases. Reported are the unadjusted (left) and the adjusted (right) Rand Index for each pair of folds.

| RI | Fold 2 | Fold 3 | Fold 4 | Fold 5 | ARI | Fold 2 | Fold 3 | Fold 4 | Fold 5 |
| --- | --- | --- | --- | --- | --- | --- | --- | --- | --- |
| Fold 1 | 0.84 | 0.81 | 0.85 | 0.83 | Fold 1 | 0.34 | 0.21 | 0.28 | 0.26 |
| Fold 2 |  | 0.83 | 0.88 | 0.82 | Fold 2 |  | 0.36 | 0.51 | 0.29 |
| Fold 3 |  |  | 0.84 | 0.83 | Fold 3 |  |  | 0.31 | 0.30 |
| Fold 4 |  |  |  | 0.83 | Fold 4 |  |  |  | 0.26 |

a. Unadjusted Rand Index (RI)
b. Adjusted Rand Index (ARI)

## 4 Discussion

The results of this study indicate that hypertensive patients can be separated into distinct complication risk subgroups defined by their protein expression profiles. These molecular signatures provide a biological basis for the clinical heterogeneity observed in hypertension and its complications, which could enable implementation of risk and mechanism-based precision medicine across clinical populations.

Addressing these subgroups individually offers several potential advantages over treating hypertension with a single uniform care pathway. Stratification could facilitate earlier identification of patients at high risk of cardiovascular and renal complications, enabling prioritization of targeted lifestyle interventions and/or more intensive weight loss and blood pressure control treatment if required. It could allow for management primarily by lifestyle advice in lower-risk patients, thereby improving the overall cost-effectiveness of hypertension management and reduction of over-medication. Finally, mechanistic investigation of the proteins driving complication development may reveal therapeutic targets – either novel or with potential to be modulated by existing drugs (i.e., drug repurposing) – offering opportunities to mitigate vascular and renal injury within specific patient subgroups where existing drugs are less effective.

### 4.1 Cluster Interpretation

The biological interpretation of the major protein expression clusters identified in patients diagnosed with hypertension provides insight into the molecular heterogeneity within the disease which is reflected in associated cardiovascular and renal complication profiles.

Notably, three clusters were associated with significant broad enrichment for these complications (clusters 4, 8 and 10), whereas three others appeared to have lower prevalence of complications (clusters 2, 6 and 9). Each cluster displayed distinct protein expression signatures that may help explain the diverse clinical outcomes observed among hypertensive patients.

#### 4.1.1 Complication-enriched Clusters

Three complication-associated clusters (clusters 4, 8, and 10) showed elevated expression of REN relative to controls, resembling the previously described high-renin hypertension subgroup characterized by increased plasma renin activity (PRA) [25]. Renin is an aspartyl protease that catalyzes the conversion of angiotensinogen to angiotensin I – the rate-limiting step of the renin–angiotensin–aldosterone system (RAAS) – thereby regulating blood pressure and sodium homeostasis [57, 58]. Although elevated renin protein levels generally lead to increased PRA, this relationship can be modulated by both biological and pharmacological factors [59].

Patients with high renin activity are at greater risk of cardiovascular and renal injury [60, 61], consistent with the complication profiles of the three clusters. Furthermore, RAAS overactivation, a downstream consequence of increased REN activity, has been implicated in the development of hypertension-associated atrial fibrillation [26, 62], a complication specifically enriched in cluster 4.

Expression of HAVCR1 was also increased in the complication-enriched clusters 4, 8 and 10, relative to controls. HAVCR1 encodes kidney injury molecule-1 (KIM-1), a phosphatidylserine receptor induced in proximal tubular epithelial cells following nephrotoxic insult [63, 64]. KIM-1 has been proposed as a biomarker of renal injury and disease severity, particularly in chronic kidney disease (CKD) and acute tubular necrosis [65–67]. In addition, HAVCR1 variants have been associated with CAD and stroke [68]. These findings further align with the enrichment of cardiovascular and renal complications in clusters 4, 8, and 10.

Similarly, RTN4R protein levels were also elevated in the three complication-enriched groups. RTN4R encodes the Nogo receptor, which binds RTN4 (Nogo-A), a negative regulator of angiogenesis and vascular network remodeling in the postnatal brain [69, 70]. Inhibition of this pathway promotes vascular repair and enhances vessel network formation after stroke [70].

Expression of RTN4R in clusters 8 and 10 may therefore reflect impaired vascular remodeling associated with ischemic injury and could help explain the enrichment for myocardial infarction (MI) for those clusters. Supporting this interpretation, RTN4 has been implicated in post-MI remodeling and recovery [71].

In contrast to these shared patterns, protein expression also diverged across the complication-associated clusters. Expression of PTPRB was modestly reduced in cluster 4 compared with controls, whereas it was increased in clusters 8 and 10, with a greater effect size in the latter.

PTPRB encodes a vascular endothelial protein tyrosine phosphatase (VE-PTP) involved in blood-vessel remodeling and angiogenesis via regulation of Tie-2 activity [72, 73]. Elevated PTPRB has been implicated in ischemia–reperfusion injury and diabetic kidney disease, where its inhibition confers renal protection [74, 75]. These observations suggest that PTPRB expression helps distinguish among the complication-associated clusters and may contribute an additional molecular signature related to susceptibility to hypertensive complications.

Taken together, the combined up-regulation of REN, HAVCR1, and RTN4R points to a protein profile indicative of heightened vascular and renal vulnerability, consistent with the clinical phenotypes of the complication-enriched clusters. At the same time, differences in individual proteins such as PTPRB highlight distinct molecular signatures across these clusters and might suggest heterogeneous underlying biology.

We observed a relationship between age and incidence of complications. This pattern is well-known [76]. However, the age differences across clusters were relatively modest (up to 3 years between low- and high-risk clusters), and the cluster with the oldest age distribution (cluster 5) did not exhibit the most severe complication profile among the age-enriched groups. This suggests that age is not the main driver behind the differences in complication prevalence between clusters.

Similarly, several clusters demonstrated sex enrichment, but no consistent trend emerged linking either sex to higher- or lower-risk phenotypes. This aligns with previous findings that sex typically acts as a secondary stratifier, contributing to patient differentiation primarily in conjunction with other factors such as BMI or metabolic status [77–79]. These interactions may warrant further investigation in future studies.

#### 4.1.2 Clusters with Lower Prevalence of Complications

In contrast to the complication-enriched clusters, expression of REN and HAVCR1 in the low-risk complication clusters was similar to those of controls. This pattern was consistent across clusters 2, 6, and 9, and the absence of up-regulation aligns with their lower risk of developing complications. By contrast, the expression of the other key proteins – PLAT, PTPRB and RTNR4 – varied among the “protected” clusters.

PLAT levels were consistently up-regulated in all clusters relative to controls, but the extent of this increase varied within the protected clusters. In clusters 2 and 6 the effect size was modest, whereas cluster 9 showed a larger increase. Altered PLAT expression and release have previously been implicated in hypertension, although the directionality and underlying mechanisms remain unclear [80, 81] potentially because its importance for hypertension varies by cluster as suggested by our results. Recent Mendelian randomization studies further suggest that PLAT may mediate a causal pathway linking smoking to essential hypertension [82], potentially connecting environmental exposures to molecular dysregulation within different hypertensive patient subgroups.

Expression of PTPRB also differentiates the low-risk clusters. Cluster 9 showed a stronger up-regulation of PTPRB than cluster 2, whereas cluster 6 exhibited a modest down-regulation. Given PTPRB’s role in promoting endothelial activation and renal injury, its differential expression across these clusters may reflect distinct protective molecular phenotypes, consistent with their differing clinical association profiles.

### 4.2 Analysis Limitations and Future Work

To obtain the results presented in this paper, we developed a proteomics-based patient stratification pipeline based on analyzing the structure of a machine learning model trained to predict existing hypertension. In this section we discuss limitations of data sources and directions for future work and improvement of the pipeline, as well as integration of multi-omic data.

The analysis presented in this paper is affected by data availability and quality. UKB proteomics data is available for approximately 10% of UKB participants, presenting challenges associated with low sample size such as reduced robustness of the detected clusters and low statistical power, as well as ancestry bias as shown in Supplementary Data, Figure S1.

Hypertension is a superficially simple phenotype, but one for which obtaining a consistent detailed clinical diagnosis sufficient to more accurately sub-phenotype patients is inherently challenging [83]. In addition, the analysis was performed on the basis of a single proteomic profile at UKB Instance 0, which does not allow for analysis of stability or treatment and comorbidity responses within individual patients.

Additionally, as hypertension is a common phenotype, the dataset used suffers from a shortage of controls. Only 16,047 hypertension controls were available, 13,016 of which had to be included in the dataset to obtain the desired 1:2 case:control ratio, limiting our ability to perform age- and sex-matching between cases and controls and thus potentially introducing covariate bias.

We expect the scope of these limitations to decrease considerably with the release of additional UKB proteomics data scheduled for 2027 [84]. As proteomics data becomes more widely available, we also expect to be able to carry out external validation of the reported findings that would increase the impact of the study by ensuring robustness of results across multiple datasets with diverse ancestry cohorts.

The analysis is further complicated by the difficulty of identifying treatment naïve patients – i.e., those whose proteomic profiles have not been altered by prescribed drugs. Drugs (especially those acting on the main disease mechanisms) often alter protein expression levels and therefore represent an important confounding factor to consider when performing proteomic-based patient stratification.

For example, during initial investigations, we found that expression of angiotensin-converting enzyme (ACE) was one key differentiator between cases and controls and between clusters that were enriched for complications versus protected clusters. However, altered expression of ACE protein acts as a differentiator between hypertension cases and controls primarily because it is strongly correlated with whether or not a person is taking an ACE inhibitor, the most common drug treatment for hypertension [35, 85].

Indeed, as expected, we identified a highly significant enrichment of patients taking ACE inhibitors in clusters enriched for complications vs. protected clusters. This suggests that the differences in ACE expression originally used to define clusters partly reflected treatment rather than biological subtypes of hypertension and therefore offered limited utility for precision medicine.

After removing patients who were taking ACE inhibitors from the dataset, the model no longer assigned high importance to ACE expression for case/control classification, or used it for clustering, confirming that a portion of our original (unreported) results were likely an artifact of differences in treatment.

This result highlights an important need for careful study designs tailored to individual phenotypes when using proteomic data to stratify patients, rather than an inflexible approach that uses the same dataset to model hundreds of phenotypes. Nonetheless, it is possible that some protein expression features used to define clusters in our current model may reflect other less common medicines that were not considered in our study design.

To address the potential for confounding factors in blood plasma proteomics, our methodological approach was focused strongly on controlling for age and sex. Controlling for covariates via incorporation in a linear model is a well-established practice in scientific literature [86–88], but equivalent approaches for non-linear models are understudied. We decided against using more elaborate deconfounding methods such as DoubleML [88], an approach which involves regressing confounders out of independent and dependent variables, since they involve additional hyperparameter choices and hinder interpretability.

Another major focus of the methods employed in this paper is replicability. We measured the stability of stratification results across training folds, indicating the extent to which they may vary if the stratification models were trained under different conditions. A few factors affect the unadjusted and adjusted Rand Index as reported in Table 4: the quality of training data, model selection, sensitivity of segregating proteins to model selection (including its training data), dimensionality reduction, and clustering algorithm output.

An artifact of machine learning models is that, regardless of the choice of model parameters, they can produce equally accurate predictions in two different ways that are equally valid. Similarly, selecting a set of “segregating proteins” is not a task for which a ground truth is available. Indeed, we observe that different sets of segregating proteins produce stratifications that share general patterns of cluster enrichment versus cluster proteomic profile.

Ultimately, patient stratification is extremely difficult without a ground truth, so among two different solutions, one is not necessarily “wrong”. Noise introduced by technical steps such as dimensionality reduction and algorithmic clustering has been included in the end-to-end stratification maps we tested for stability.

The ARI values reported in Table 4b measure the similarity between two stratifications on a scale from random to perfect, so an ARI of 0.5 would already be classed as satisfactory as it represents a level of agreement akin to an AUROC score of 0.75 (since the AUROC score of a random classifier is 0.5). Based on the results presented in this paper, the development of robust but not artificially deterministic stratification algorithms aiming at an ARI of 0.75 is a high-priority research objective of future work.

We expect increasing the sample size and depth and quality of phenotyping to be the most effective way of achieving this. The scheduled release of proteomic data for all UKB participants in 2027 should facilitate development of more stable machine learning models for hypertension. Additionally, we expect that applying this approach to other health conditions with higher quality phenotyping data will result in higher ARI stability scores.

The clustering algorithm used in this paper is spectral clustering [48, 49], and was chosen due to its ability to detect non-convex clusters of various shapes. The number of clusters is a free parameter of this algorithm, and for this paper this was chosen via visual inspection of the available data points based on their shape and appearance, and to ensure a minimum cluster size. Approaches such as the elbow method [49] and eigengap method [89], which is specific to spectral clustering, can be employed in future work to determine the optimal number of clusters automatically.

Replacing manual inspection with those automatic criteria would fully automate the stratification pipeline and make it reproducible, reducing potential human bias in the choice of UMAP hyperparameters and number of clusters. Preliminary analyses with automatic selection of the number of clusters indicate that it yields distinct enriched and protected patient groups with clustering quality metrics comparable to the manual alternative, providing a coherent basis for successive biological interpretation.

In addition, a final global stratification step can aggregate the information across all cross-validation folds to derive a single patient clustering. Rather than inspecting and comparing fold-specific clusterings, this global stratification would offer a unified view of patient subgroups and their biological interpretation. In this framework, patients are first clustered within each fold and common segregating proteins (see 2.2.4 Methods for Stratification) are combined to identify a set of globally informative features. Protein contributions for cases are then aggregated across folds on this common feature set, and a single clustering is learned on the resulting representation.

While plasma proteomics is highly predictive of common phenotypes [19], it cannot directly comment on etiology or causality as associations between proteome and phenotype can be both due to upstream and downstream effects. A fruitful direction of further work addressing this limitation could be to employ causal approaches, such as Mendelian randomization [90], or genetically predicted protein levels (GPPLs) incorporating genomic data to substantiate causal links between protein expression and phenotype [91].

We believe that the most productive approach that will generate more clinical utility is to combine the causal mechanistic stratification enabled by combinatorial genetic analysis [92] with proteomic clustering. Combinatorial analysis has been shown to generate more genetic associations than GWAS with better reproducibility across diverse populations in many complex, chronic diseases [93–97]. Crucially it has also been shown to be able to identify subgroups with shared mechanistic etiology that correlate with clinical symptoms [93, 94]. The proteomic profiles of patients within these mechanistic clusters can be expected to be more similar to each other than those between different clusters.

Given that many complex diseases have multiple distinct mechanistic etiologies [93–97], we would expect that each of these would affect the proteomes of people in that cluster in different ways. In these diseases many people have more than one mechanistic driver of their disease so may present with a mix of features. To maximize the predictive value of a proteomic (or other multi-omic) biomarkers, this makes it important to consider genetic / mechanistic clustering before running clustering and ‘omics data to avoid conflation of expression / epigenetic signals arising for different subsets of patients.

This may help further deconvolute causal mechanisms and their impact on proteomic signals, more accurately assess risk (and resilience), and improve the precision of treatment selection. This would potentially overcome some of the regulatory and clinical issues associated with the proposed use of single sample proteomics [98] and root the signal squarely in causal biology.

## 5 Conclusion

We applied a novel patient stratification pipeline to proteomics data from a cohort of hypertensive patients to identify clusters of hypertension cases associated with differing prevalence of cardiovascular and renal complications. The main novelty of the approach lies in exploiting the contributions of individual proteins to the output of a tree-based risk model for multiple clinically relevant phenotypes of interest. Unlike using raw data for clustering, using the structure of a tree-based model leverages combinations of protein expression values that inform the model, which naturally separates its input data into groups. As a result, the hypertension patient clusters presented in this paper show a greater degree of separation compared to alternative proteomic-based stratification approaches [32, 34].

We hypothesize that the hypertension clusters identified in this study may represent distinct mechanistic subtypes. This appears to be supported by their distinct phenotype enrichments. These can potentially be used by researchers to more efficiently focus studies on subgroups of hypertension patients who have similar disease etiology, identify precision medicine treatments that are most effective for each subgroup, and use mechanistic stratification approaches to inform the design and improve the likelihood of success of clinical drug trials.

## Data and Code Availability

Research data including raw or processed data files, software, code, models, algorithms, protocols and methods is proprietary and may be shared upon reasonable request and under an appropriate data access agreement.

## CRediT Authorship Contribution Statement

**YP:** Conceptualization, Data curation, Formal analysis, Investigation, Methodology, Project administration, Software, Supervision, Validation, Visualization, Writing – original draft, Writing – review and editing. **ARM:** Investigation, Writing – original draft. **SA**: Conceptualization, Formal analysis, Investigation, Methodology, Software, Visualization, Writing – original draft. **DK:** Conceptualization, Formal analysis, Investigation, Methodology, Software, Supervision, Writing – original draft. **MP**: Conceptualization, Project administration, Supervision. **JS:** Conceptualization, Project administration, Supervision, Writing – review and editing. **MAS:** Project administration, Supervision, Writing – review & editing. **SG:** Data access, Writing – review and editing.

## Funding Sources

This research did not receive any specific grant from funding agencies in the public, commercial, or not-for-profit sectors.

## Conflict of Interest

SG is a shareholder of and has assigned patent rights to PrecisionLife Ltd. SG is an Editorial Board member of AILSCI.

## Data Availability

Research data including raw or processed data files may be shared upon reasonable request and under an appropriate data access agreement.

## Acknowledgements

We thank Maia Cooper for conducting an initial literature review for this manuscript.

This research has been conducted using the UK Biobank Resource under Application Number 44288. UK Biobank has approval from the North West Multi-centre Research Ethics Committee (MREC) as a Research Tissue Bank (RTB) approval, and researchers do not require separate ethical clearance and can operate under this RTB approval. This work uses data provided by patients and collected by the NHS as part of their care and support, for which we are also grateful.

## Notes

### Funding Statement

This research did not receive any funding.

### Author Declarations

This research has been conducted using the UK Biobank Resource under Application Number 44288. UK Biobank has approval from the North West Multi-centre Research Ethics Committee (MREC) as a Research Tissue Bank (RTB) approval, and researchers do not require separate ethical clearance and can operate under this RTB approval.

