## Supplementary Methods and Data for "Proteomics Reveal Clusters of Hypertension Cases Associated with Differing Prevalence of Cardiovascular and Renal Complications"

**Supplementary Data**

**Study Design**

**Table S1.** Summary of study design and list of relevant complications for our hypertension dataset. Controls were down-sampled to a 1:2 case:control ratio as described in Methods, Study Design.

| **Cases** | - I10 in data-field 41270 and date I10 first reported (field 131286) earlier than proteomics sample (field 53, instance 0) - Not on ACE inhibitors (field 20003, excluded codes in - Table S2 below)   Yielding 7,086 cases |
| --- | --- |
| **Controls** | - No I10-15 in data-field 41270 (at any time) - BP between 90/60 and 135/85 from automated reading (fields 4079 and 4080, for each averaging both array entries) and - No BP medication (data-field 6177, data-coding 100625 value 2)   Yielding 13,016 controls |
| **Complications (sourced from data-field 41270)** | - myocardial infarction I21-22 - atrial fibrillation I48 - chronic ischaemic heart disease I25 - heart failure I11, I50 - renal failure I12, N17, N18 - cardiac arrest I46 - stroke I60-64 - cardiomyopathy I42 (not timed) |

**Table S2.** Medication codes for ACE inhibitors excluded from the cohort of hypertension cases.

| Code | Medication name |
| --- | --- |
| 1140860696 | lisinopril |
| 1140860728 | quinapril |
| 1140860738 | quinalapril+hydrochlorothiazide 10mg/12.5mg tablet |
| 1140860750 | captopril |
| 1140860752 | acepril 12.5mg tablet |
| 1140860764 | captopril+hydrochlorothiazide 25mg/12.5mg tablet |
| 1140860790 | enalapril maleate+hydrochlorothiazide 20mg/12.5mg tablet |
| 1140860806 | ramipril |
| 1140860882 | cilazapril |
| 1140860904 | trandolapril |
| 1140864952 | lisinopril+hydrochlorothiazide 10mg/12.5mg tablet |
| 1140888552 | enalapril |
| 1140888556 | fosinopril |
| 1140888560 | perindopril |
| 1140923712 | moexipril |
| 1141151382 | hypapril 12.5mg tablet |
| 1141153328 | trandolapril+verapamil hydrochloride |
| 1141164148 | imidapril hydrochloride |
| 1141165470 | felodipine+ramipril |
| 1141167822 | tensopril 12.5mg tablet |
| 1141180592 | perindopril+indapamide |

**Table S3.** Age and sex distribution of cases and controls
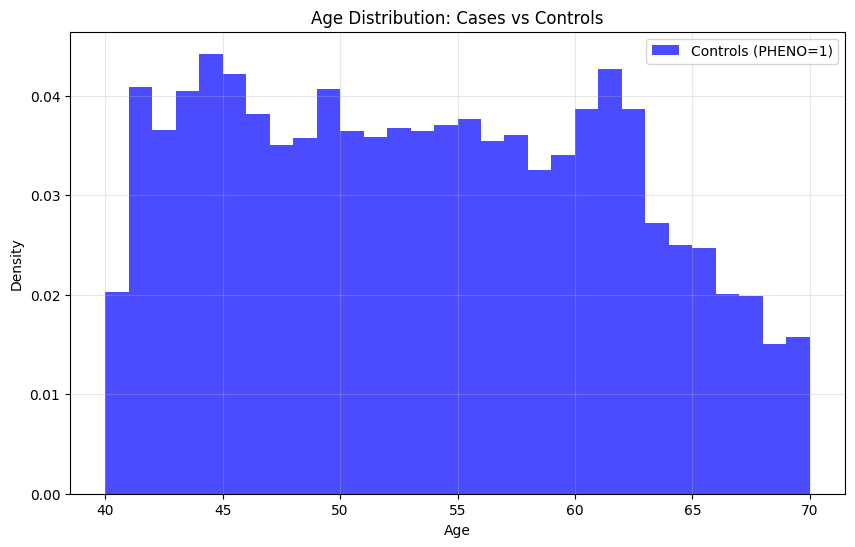

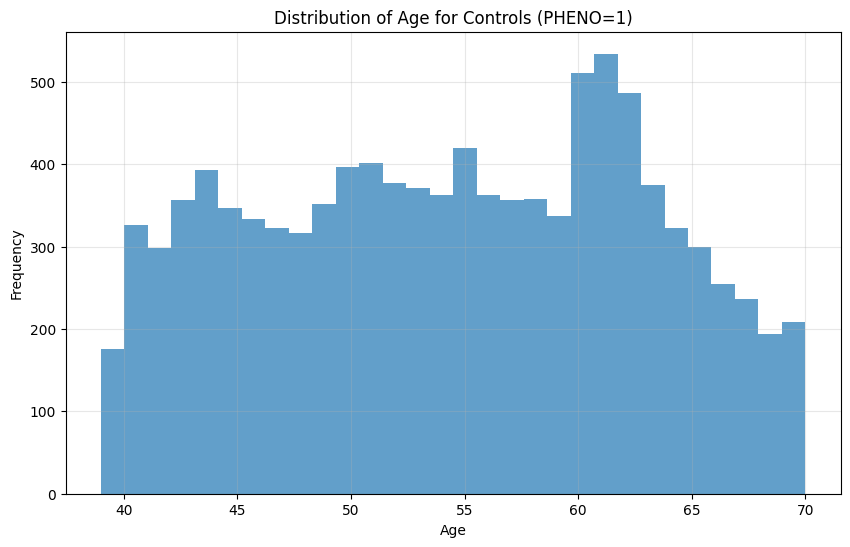
after age- and sex-matching controls to cases in a 1:2 case:control ratio. Age was sourced from Data-field 21003, at instance 0 and sex was sourced from Data-field 31.

|  | **Cases** | **Controls** |
| --- | --- | --- |
| **Sex** | 51% female | 63% female |
| **Age** | 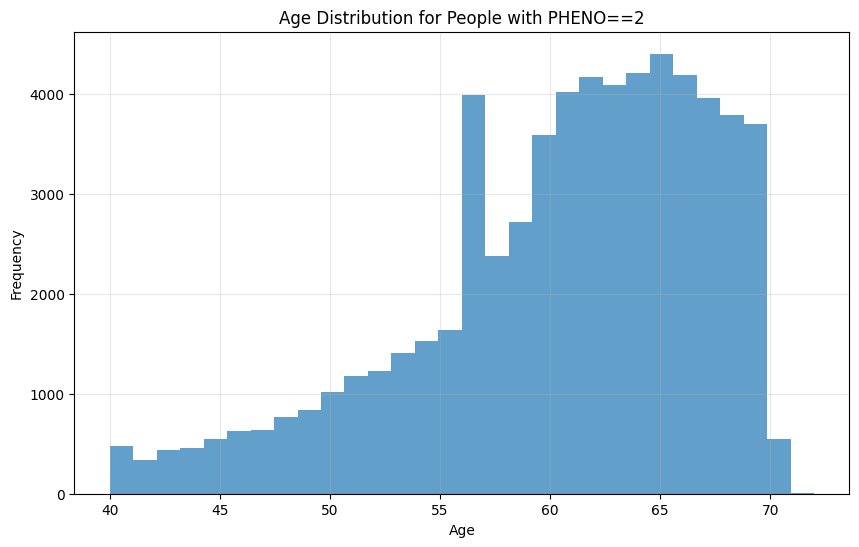 | 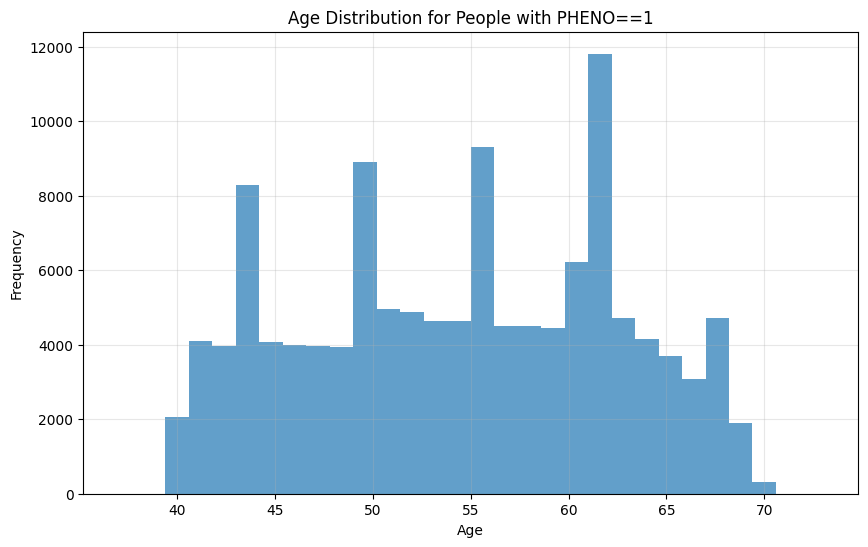 |

**Table S4.** Number of hypertension cases and controls used for training and stability analysis.

|  | **Cases** | **Controls** | **Both** |
| --- | --- | --- | --- |
| **All samples** | 7,086 | 13,016 | 20,102 |
| **Used for training** | 5,669 | 10,412 | 16,081 |
| **Used for stability analysis** | 1,417 | 2,604 | 4,021 |

|  |  |
| --- | --- |
| European | 13852 |
| Latin American 1 | 710 |
| Latin American 2 | 23 |
| African | 470 |
| South Asian | 271 |
| East Asian | 37 |
| Other | 582 |
| No data | 136 |

|  | Ethnicity |
| --- | --- |
| White: British | 13918 |
| White: Other | 574 |
| White: Irish | 397 |
| African | 213 |
| Other ethnic group | 206 |
| Indian | 186 |
| Caribbean | 178 |
| Other categories | 409 |

**Figure S1.** Genetic ancestry distribution of all samples in our hypertension dataset obtained using [GRAF-pop](https://github.com/ncbi/graf) (left) and self-reported ethnic background from UK Biobank Data-Field 21000 (right). Ethnic background categories with fewer than 100 samples were collated into “Other categories”. Definitions of the genetic ancestry categories are described in Tables 2 and 3 in [32].

**Proteins Removed During QC**

A list of 12 proteins were removed due to missingness, shown in the table below.

**Table S5**. Proteins removed due to missingness.

| **Coding** | **Protein** |
| --- | --- |
| 113 | AMY1A AMY1B AMY1C; Alpha-amylase 1A Alpha-amylase 1B Alpha-amylase 1C |
| 1173 | GLIPR1;Glioma pathogenesis-related protein 1 |
| 1569 | LEG1;Protein LEG1 homolog |
| 1889 | NPM1;Nucleophosmin |
| 1937 | OLFM4;Olfactomedin-4 |
| 1991 | PCOLCE;Procollagen C-endopeptidase enhancer 1 |
| 2182 | PSTPIP2; Proline-serine-threonine phosphatase-interacting protein 2 |
| 277 | BPIFA2;BPI fold-containing family A member 2 |
| 542 | CFH;Complement factor H |
| 709 | CST1;Cystatin-SN |
| 732 | CTSS;Cathepsin S |
| 923 | ENDOU;Poly(U)-specific endoribonuclease |

**Model Training**

**XGBoost Hyperparameter Search Space**

During hyperparameter optimization the following search space is passed to a BayesSearchCV object from the python package scikit-optimize:

```
search_space = {'max_depth': Integer(2,5),
                'reg_alpha': Real(low=0, high=150, prior='uniform'),
                'learning_rate': Real(low=0.001, high=0.1, prior='uniform'),
                'n_estimators': [100, 500, 1000, 2000]}
```

**System Specifications**

To generate the results in this paper, we used UKB-RAP’s mem2_ssd1_v2_x64 AWS instance with 64 CPU cores and 256GB RAM. More details on the compute environment can be found under [Instance Types in UKB-RAP’s online documentation pages](https://documentation.dnanexus.com/developer/api/running-analyses/instance-types). A five-fold cross-validation run took approximately 35min.

**Model Metrics**

As well as AUROC, we report mean and sample standard deviation of our models’ balanced accuracy, precision, and recall across all five folds. Note that these metrics are computed with the default decision threshold of 0.5, and can be further optimized by varying the threshold. The decision threshold doesn’t affect our stratification analyses so it has not been optimized. For the parameters used in each model, refer to Table 1. For brevity we report these metrics only for the models including age and sex as covariates.

**Table S6.** Mean and sample standard deviation of train and test model performance metrics across five folds.

|  | Train | Test |
| --- | --- | --- |
| Balanced accuracy | 0.997 ± 0.0002 | 0.84 ± 0.0067 |
| Precision | 0.998 ± 0.0003 | 0.83 ± 0.0099 |
| Recall | 0.996 ± 0.0005 | 0.79 ± 0.0093 |

**Additional Stratification Results**

**Fold 1**

*Train*

**Table S7.** Cohen's d effect sizes from Mann-Whitney U tests comparing protein expression levels between cases in each cluster and the cohort of all cases or controls. Arrows indicate the direction of regulation of protein expression. Rows highlighted in cyan indicate protected clusters, while rows highlighted in red indicate clusters enriched for complications.

| **Cluster** | **HAVCR1** | | **PLAT** | | **PTPRB** | | **REN** | | **RTN4R** | |
| --- | --- | --- | --- | --- | --- | --- | --- | --- | --- | --- |
|  | vs controls | vs  cases | vs controls | vs  cases | vs controls | vs  cases | vs controls | vs  cases | vs controls | vs  cases |
| **1** | ▲1.542 | ▲0.617 | ▲0.800 | ▲0.054 | ▲0.553 | ▲0.014 | ▲0.346 | ▼-0.203 | ▲0.671 | ▲0.073 |
| **2** | ▼-0.158 | ▼-1.215 | ▲0.501 | ▼-0.299 | ▲0.429 | ▼-0.117 | ▲0.101 | ▼-0.365 | ▲0.389 | ▼-0.254 |
| **3** | ▲1.506 | ▲0.529 | ▲0.656 | ▼-0.107 | ▼-0.556 | ▼-1.288 | ▲0.366 | ▼-0.170 | ▲0.580 | ▼-0.032 |
| **4** | ▲1.672 | ▲0.643 | ▲0.555 | ▼-0.200 | ▼-0.484 | ▼-1.043 | ▲1.690 | ▲0.698 | ▲0.872 | ▲0.251 |
| **5** | ▼-0.114 | ▼-0.982 | ▲0.572 | ▼-0.191 | ▲0.387 | ▼-0.141 | ▲0.719 | ▲0.112 | ▲0.384 | ▼-0.231 |
| **6** | ▼-0.231 | ▼-1.114 | ▲0.417 | ▼-0.358 | ▼-0.616 | ▼-1.310 | ▲0.190 | ▼-0.274 | ▲0.324 | ▼-0.295 |
| **7** | ▲1.650 | ▲0.691 | ▲1.081 | ▲0.366 | ▲1.797 | ▲1.637 | ▲0.379 | ▼-0.167 | ▲0.765 | ▲0.164 |
| **8** | ▲1.939 | ▲0.952 | ▲0.943 | ▲0.198 | ▲0.578 | ▲0.050 | ▲1.989 | ▲1.011 | ▲0.947 | ▲0.341 |
| **9** | ▼-0.057 | ▼-0.893 | ▲1.267 | ▲0.520 | ▲1.637 | ▲1.185 | ▲0.310 | ▼-0.193 | ▲0.576 | ▼-0.035 |
| **10** | ▲2.016 | ▲0.965 | ▲1.082 | ▲0.334 | ▲1.715 | ▲1.251 | ▲2.102 | ▲0.951 | ▲0.999 | ▲0.380 |

**Table S8.** *P*-values for protein expression differences between cases in each cluster and controls. Bold denotes *p*-values that were significant at the *p* < 0.05 level after applying Benjamini-Hochberg FDR correction.

| Cluster | HAVCR1 | PLAT | PTPRB | REN | RTN4R |
| --- | --- | --- | --- | --- | --- |
| 1 | **0.00E+00** | **1.38E-106** | **4.33E-62** | **6.78E-24** | **5.87E-74** |
| 2 | **1.21E-04** | **1.92E-47** | **4.02E-37** | **2.75E-02** | **2.71E-27** |
| 3 | **1.46E-189** | **1.49E-44** | **2.44E-41** | **1.37E-15** | **1.99E-33** |
| 4 | **6.33E-82** | **1.19E-13** | **1.72E-15** | **2.25E-17** | **1.11E-27** |
| 5 | 9.95E-02 | **5.69E-26** | **9.01E-12** | **6.71E-03** | **9.06E-13** |
| 6 | **2.41E-05** | **6.33E-17** | **3.33E-41** | **1.76E-05** | **7.19E-11** |
| 7 | **2.69E-238** | **2.49E-115** | **6.77E-276** | **5.57E-20** | **4.43E-65** |
| 8 | **1.29E-158** | **3.48E-59** | **1.60E-29** | **1.08E-55** | **2.20E-53** |
| 9 | 8.92E-01 | **1.87E-71** | **2.52E-123** | **1.69E-06** | **1.05E-18** |
| 10 | **4.86E-100** | **3.73E-39** | **1.24E-100** | **1.70E-37** | **1.09E-36** |

**Table S9.** *P*-values for protein expression differences between cases in each cluster and all remaining cases. Bold denotes *p*-values that were significant at the *p* < 0.05 level after applying Benjamini-Hochberg FDR correction.

| Cluster | HAVCR1 | PLAT | PTPRB | REN | RTN4R |
| --- | --- | --- | --- | --- | --- |
| 1 | **4.55E-91** | 8.02E-02 | 2.55E-01 | **2.93E-03** | **3.82E-02** |
| 2 | **6.04E-233** | **3.30E-13** | **3.13E-02** | **1.45E-18** | **1.02E-11** |
| 3 | **4.01E-43** | **2.26E-02** | **7.93E-157** | 6.73E-02 | 6.14E-01 |
| 4 | **1.38E-22** | **1.37E-02** | **2.47E-54** | **1.72E-11** | **1.62E-03** |
| 5 | **8.05E-89** | **2.21E-04** | 4.28E-02 | 6.79E-01 | **1.83E-05** |
| 6 | **7.23E-113** | **2.09E-13** | **1.66E-133** | **8.98E-06** | **1.60E-08** |
| 7 | **8.98E-73** | **2.88E-16** | **7.52E-229** | 8.08E-02 | **4.24E-05** |
| 8 | **3.90E-60** | **3.23E-04** | 1.85E-01 | **9.81E-41** | **6.92E-09** |
| 9 | **1.01E-54** | **2.78E-16** | **2.41E-83** | 6.36E-02 | 8.49E-01 |
| 10 | **2.62E-41** | **8.34E-05** | **4.70E-69** | **3.21E-26** | **6.34E-08** |

*Test*

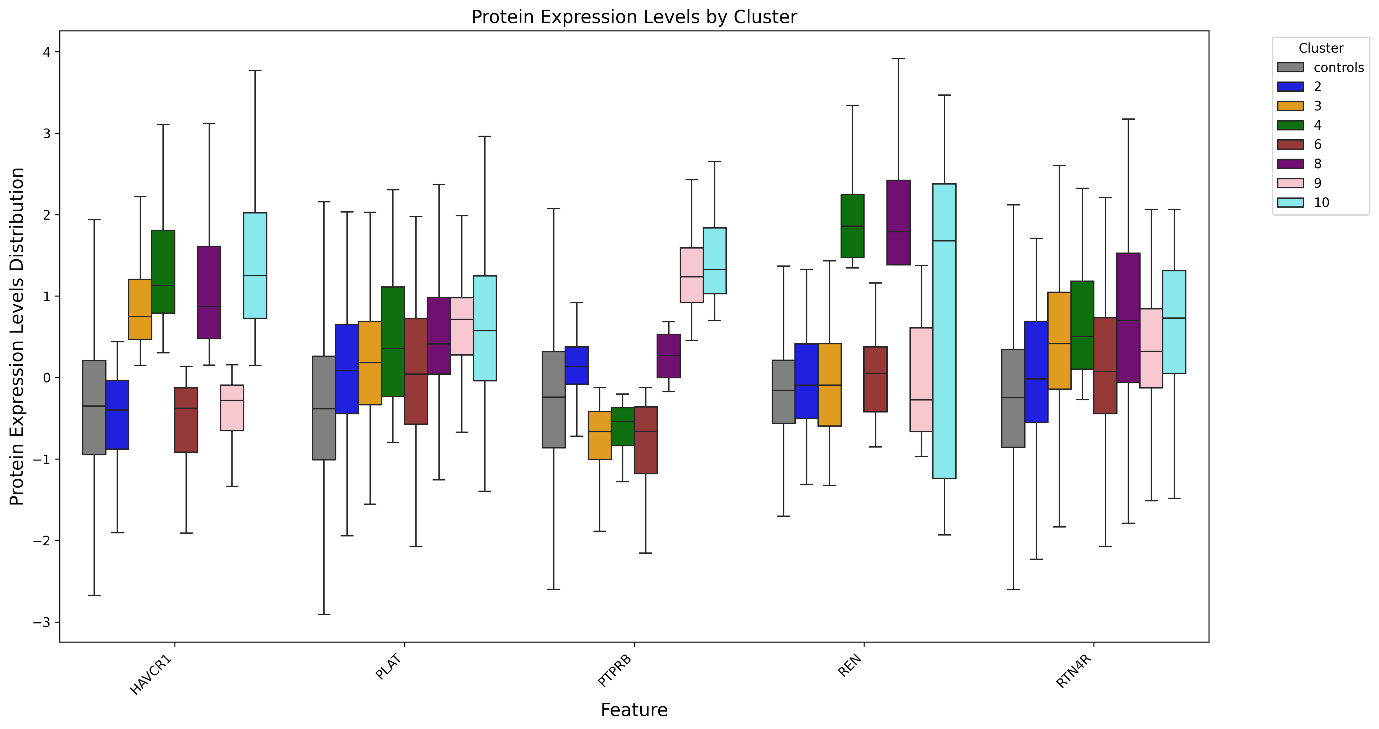

**Figure S2.** Differential expression of separating proteins among hypertension cases

**Table S10**. Cohen's d effect sizes from Mann-Whitney U tests comparing protein expression levels between cases in each cluster and controls. Upward-pointing arrows indicate higher expression in cases; downward-pointing arrows indicate higher expression in controls.

| Cluster | HAVCR1 | PLAT | PTPRB | REN | RTN4R |
| --- | --- | --- | --- | --- | --- |
| 1 | ▲0.154 | ▲0.820 | ▲0.547 | ▲0.273 | ▲0.580 |
| 2 | ▼-0.023 | ▲0.465 | ▲0.399 | ▲0.130 | ▲0.253 |
| 3 | ▲0.139 | ▲0.562 | ▼-0.569 | ▲0.118 | ▲0.709 |
| 4 | ▲1.920 | ▲0.865 | ▼-0.426 | ▲0.142 | ▲0.999 |
| 5 | ▼-0.133 | ▲0.569 | ▲0.405 | ▲0.125 | ▲0.526 |
| 6 | ▼-0.307 | ▲0.445 | ▼-0.643 | ▲0.211 | ▲0.353 |
| 7 | ▲1.780 | ▲1.060 | ▲1.78 | ▲0.279 | ▲0.573 |
| 8 | ▲1.720 | ▲0.941 | ▲0.526 | ▲1.920 | ▲1.010 |
| 9 | ▼-0.123 | ▲1.060 | ▲1.670 | ▲0.099 | ▲0.610 |
| 10 | ▲1.950 | ▲1.070 | ▲1.800 | ▲0.161 | ▲0.874 |

**Table S11.** *P*-values for protein expression differences between cases in each cluster and controls. Bold denotes *p*-values that were significant at the *p* < 0.05 level after applying Benjamini-Hochberg FDR correction.

| Cluster | HAVCR1 | PLAT | PTPRB | REN | RTN4R |
| --- | --- | --- | --- | --- | --- |
| 1 | **5.03E-83** | **3.29E-29** | **3.02E-16** | **1.25E-04** | **9.12E-16** |
| 2 | **6.31E-03** | **1.25E-10** | **4.22E-09** | **4.49E-02** | **2.63E-04** |
| 3 | **5.01E-51** | **1.80E-11** | **2.41E-13** | 1.60E-01 | **1.18E-14** |
| 4 | **2.19E-20** | **2.60E-07** | **1.01E-03** | **5.25E-10** | **1.46E-09** |
| 5 | 3.33E-01 | **2.85E-08** | **1.46E-04** | **8.25E-07** | **4.07E-08** |
| 6 | **9.27E-03** | **1.70E-05** | **5.78E-14** | **5.00E-03** | **5.44E-05** |
| 7 | **1.65E-60** | **2.89E-26** | **5.24E-64** | **8.50E-04** | **1.76E-11** |
| 8 | **1.21E-38** | **1.67E-16** | **4.04E-08** | **2.80E-16** | **1.84E-16** |
| 9 | 5.69E-01 | **1.22E-13** | **3.62E-26** | 8.16E-01 | **3.75E-06** |
| 10 | **7.02E-25** | **5.80E-11** | **9.95E-27** | **1.41E-06** | **3.52E-10** |

**Table S12.** Cohen's d effect sizes from Mann-Whitney U tests comparing protein expression levels between cases in each cluster and all remaining cases. Upward-pointing arrows indicate higher expression in the corresponding cluster; downward-pointing arrows indicate higher expression in remaining clusters.

| Cluster | HAVCR1 | PLAT | PTPRB | REN | RTN4R |
| --- | --- | --- | --- | --- | --- |
| 1 | ▲0.662 | ▲0.109 | ▲0.075 | ▼-0.250 | ▲0.004 |
| 2 | ▼-1.206 | ▼-0.318 | ▼-0.082 | ▼-0.343 | ▼-0.386 |
| 3 | ▲0.441 | ▼-0.193 | ▼-1.241 | ▼-0.330 | ▲0.150 |
| 4 | ▲0.878 | ▲0.132 | ▼-0.898 | ▲0.993 | ▲0.428 |
| 5 | ▼-0.952 | ▼-0.170 | ▼-0.060 | ▲0.521 | ▼-0.058 |
| 6 | ▼-1.157 | ▼-0.311 | ▼-1.297 | ▼-0.264 | ▼-0.248 |
| 7 | ▲0.835 | ▲0.368 | ▲1.660 | ▼-0.224 | ▼-0.007 |
| 8 | ▲0.764 | ▲0.220 | ▲0.058 | ▲1.026 | ▲0.471 |
| 9 | ▼-0.884 | ▲0.330 | ▲1.261 | ▼-0.315 | ▲0.030 |
| 10 | ▲0.917 | ▲0.344 | ▲1.398 | ▲0.672 | ▲0.305 |

**Table S13.** *P*-values for protein expression differences between cases in each cluster and all remaining cases. Bold denotes *p*-values that were significant at the *p* < 0.05 level after applying Benjamini-Hochberg FDR correction.

| Cluster | HAVCR1 | PLAT | PTPRB | REN | RTN4R |
| --- | --- | --- | --- | --- | --- |
| 1 | **1.57E-24** | 8.89E-02 | 5.62E-02 | **2.49E-02** | 5.99E-01 |
| 2 | **1.86E-55** | **7.69E-05** | 6.92E-01 | **4.32E-04** | **3.77E-06** |
| 3 | **2.56E-11** | **2.58E-02** | **9.12E-42** | **2.73E-03** | 1.14E-01 |
| 4 | **1.05E-08** | 5.05E-01 | **1.84E-10** | **5.34E-07** | **1.18E-02** |
| 5 | **3.08E-25** | 2.23E-01 | 8.25E-01 | **3.47E-04** | 8.17E-01 |
| 6 | **8.98E-34** | **5.91E-04** | **7.48E-38** | 7.19E-02 | **1.09E-02** |
| 7 | **2.37E-24** | **8.12E-05** | **1.48E-53** | 1.24E-01 | 7.76E-01 |
| 8 | **1.46E-11** | 7.72E-02 | 2.12E-01 | **6.96E-12** | **1.71E-04** |
| 9 | **1.48E-12** | **4.84E-03** | **1.33E-18** | 4.21E-02 | 6.47E-01 |
| 10 | **1.44E-10** | 4.04E-02 | **2.34E-20** | **2.35E-04** | **6.35E-03** |

**Table S14.** Fisher's exact test results on test data for complication enrichments among hypertension cases in each cluster. Only complications that were statistically significant in train data according to the criteria in Enrichment analysis, and significant at the *p* < 0.05 level after Benjamini-Hochberg FDR correction are reported.

| Cluster | Complication | p-value | Odds ratio |
| --- | --- | --- | --- |
| **2** | **atrial fibrillation** | **2E-03** | **0.50** |
| **2** | **renal failure** | **7E-06** | **0.37** |
| **8** | **CIHD** | **6E-04** | **2.14** |
| **8** | **heart failure** | **4E-04** | **2.79** |
| **8** | **renal failure** | **2E-06** | **3.00** |
| **10** | **renal failure** | **6E-03** | **2.28** |

**Table S15.** Wald test *p*-values and odds ratios for the logistic regression coefficient of each cluster as a predictor of each complication reported in

Table **S14** on its own and in the presence of age and sex. The *p*-values in the label, sex, and age model marked bold are significant at the *p* < 0.05 level after Benjamini-Hochberg FDR correction.

| **Cluster** | **Complication** | ***p*-value**  **(label only)** | **Odds ratio**  **(label only)** | ***p*-value**  **(label + sex + age)** | **Odds ratio**  **(label + sex + age)** |
| --- | --- | --- | --- | --- | --- |
| **2** | **atrial fibrillation** | **4E-03** | **0.52** | **3E-02** | **0.59** |
| **2** | **renal failure** | **5E-05** | **0.39** | **2E-04** | **0.42** |
| **8** | **CIHD** | **6E-04** | **2.06** | **1E-02** | **1.73** |
| **8** | **heart failure** | **2E-04** | **2.60** | **1E-03** | **2.31** |
| **8** | **renal failure** | **6E-07** | **2.86** | **1E-05** | **2.53** |
| **10** | **renal failure** | **1E-02** | **2.13** | **9E-03** | **2.19** |

**Fold 2**

*Train*

| 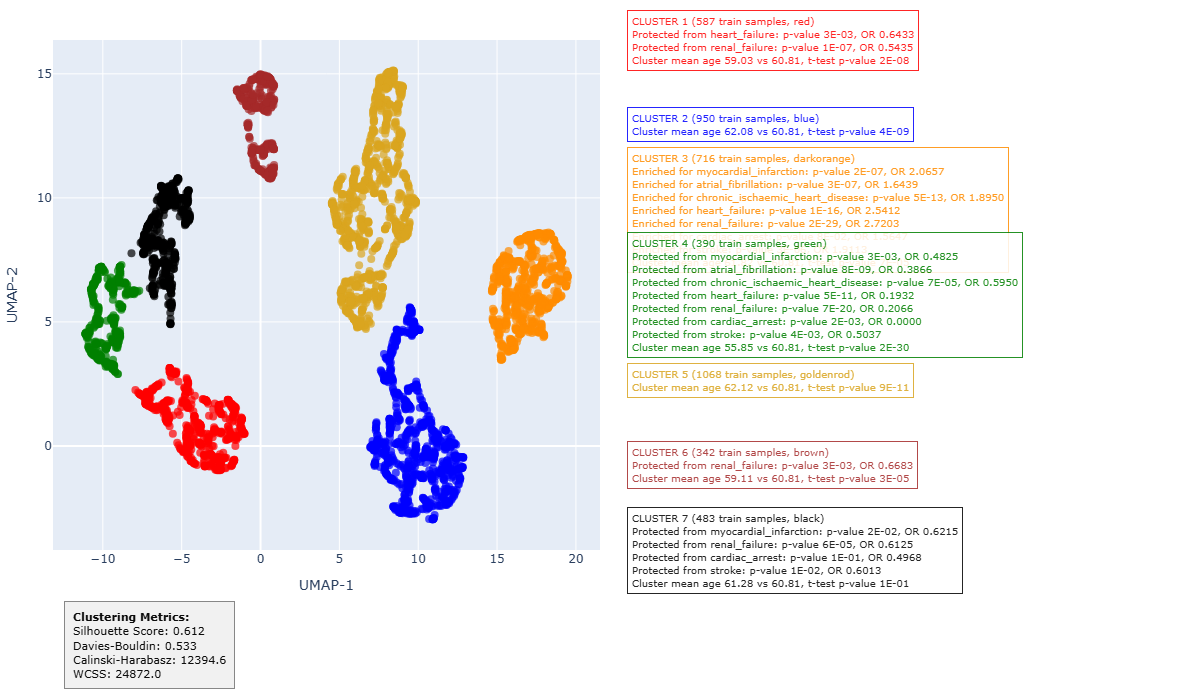 | **CLUSTER 1 (587 train samples, red)** Protected from heart failure: p-value 3E-03, OR 0.6433 Protected from renal failure: p-value 1E-07, OR 0.5435 Cluster mean age 59.03 vs 60.81, t-test p-value 2E-08 |
| --- | --- |
|  | **CLUSTER 3 (716 train samples, dark orange)** Enriched for myocardial infarction: p-value 2E-07, OR 2.0657 Enriched for atrial fibrillation: p-value 3E-07, OR 1.6439 Enriched for CIHD: p-value 5E-13, OR 1.8950 Enriched for heart failure: p-value 1E-16, OR 2.5412 Enriched for renal failure: p-value 2E-29, OR 2.7203  Enriched for stroke: p-value 3E-06, OR 1.9113 Cluster mean age 61.82 vs 60.81, t-test p-value 7E-05 |
|  | **CLUSTER 4 (390 train samples, green)** Protected from myocardial infarction: p-value 3E-03, OR 0.4825 Protected from atrial fibrillation: p-value 8E-09, OR 0.3866 Protected from CIHD: p-value 7E-05, OR 0.5950 Protected from heart failure: p-value 5E-11, OR 0.1932 Protected from renal failure: p-value 7E-20, OR 0.2066 Protected from cardiac arrest: p-value 2E-03, OR 0.0000 Protected from stroke: p-value 4E-03, OR 0.5037 Cluster mean age 55.85 vs 60.81, t-test p-value 2E-30 |
|  | **CLUSTER 6 (342 train samples, brown)** Protected from renal failure: p-value 3E-03, OR 0.6683  Enriched for cardiomyopathy: p-value 1E-02, OR 2.4964 Cluster mean age 59.11 vs 60.81, t-test p-value 3E-05 |
|  | **CLUSTER 7 (483 train samples, black)** Protected from myocardial infarction: p-value 2E-02, OR 0.6215 Protected from renal failure: p-value 6E-05, OR 0.6125 Protected from stroke: p-value 1E-02, OR 0.6013 |

**Figure S3.** Enrichments for complications, age and sex among hypertension cases from training dataset, fold 2. The silhouette score of the clustering is 0.61.

**Table S16.** Wald test *p*-values and odds ratios for the logistic regression coefficient of each cluster as a predictor of each complication reported in Figure S3 on its own and in the presence of age and sex. The *p*-values in the label, sex, and age model marked bold are significant at the *p* < 0.05 level after Benjamini-Hochberg FDR correction.

| **Cluster** | **Complication** | ***p*-value**  **(label only)** | **Odds ratio**  **(label only)** | ***p*-value**  **(label + sex + age)** | **Odds ratio**  **(label + sex + age)** |
| --- | --- | --- | --- | --- | --- |
| **1** | **heart failure** | **5E-03** | **0.65** | **4E-02** | **0.73** |
| **1** | **renal failure** | **3E-07** | **0.55** | **2E-05** | **0.61** |
| **3** | **myocardial infarction** | **1E-09** | **2.04** | **3E-08** | **1.92** |
| **3** | **atrial fibrillation** | **1E-08** | **1.64** | **7E-06** | **1.48** |
| **3** | **chronic ischaemic heart disease** | **4E-16** | **1.89** | **2E-12** | **1.75** |
| **3** | **heart failure** | **0E+00** | **2.51** | **0E+00** | **2.34** |
| **3** | **renal failure** | **0E+00** | **2.7** | **0E+00** | **2.57** |
| **3** | **stroke** | **8E-08** | **1.89** | **1E-06** | **1.79** |
| **4** | **myocardial infarction** | **1E-02** | **0.51** | **3E-02** | **0.57** |
| **4** | **atrial fibrillation** | **3E-07** | **0.4** | **2E-03** | **0.57** |
| **4** | **chronic ischaemic heart disease** | **2E-04** | **0.6** | **4E-02** | **0.75** |
| **4** | **heart failure** | **5E-07** | **0.22** | **3E-05** | **0.28** |
| **4** | **renal failure** | **1E-13** | **0.22** | **4E-10** | **0.28** |
| 4 | cardiac arrest | 7E-02 | 0.23 | 1E-01 | 0.26 |
| 4 | stroke | 1E-02 | 0.53 | 6E-02 | 0.61 |
| **6** | **renal failure** | **6E-03** | **0.67** | **4E-02** | **0.74** |
| **6** | **cardiomyopathy** | **1E-02** | **2.27** | **2E-02** | **2.19** |
| **7** | **myocardial infarction** | **4E-02** | **0.64** | **2E-02** | **0.61** |
| **7** | **renal failure** | **1E-04** | **0.62** | **3E-05** | **0.59** |
| **7** | **stroke** | **3E-02** | **0.62** | **2E-02** | **0.6** |

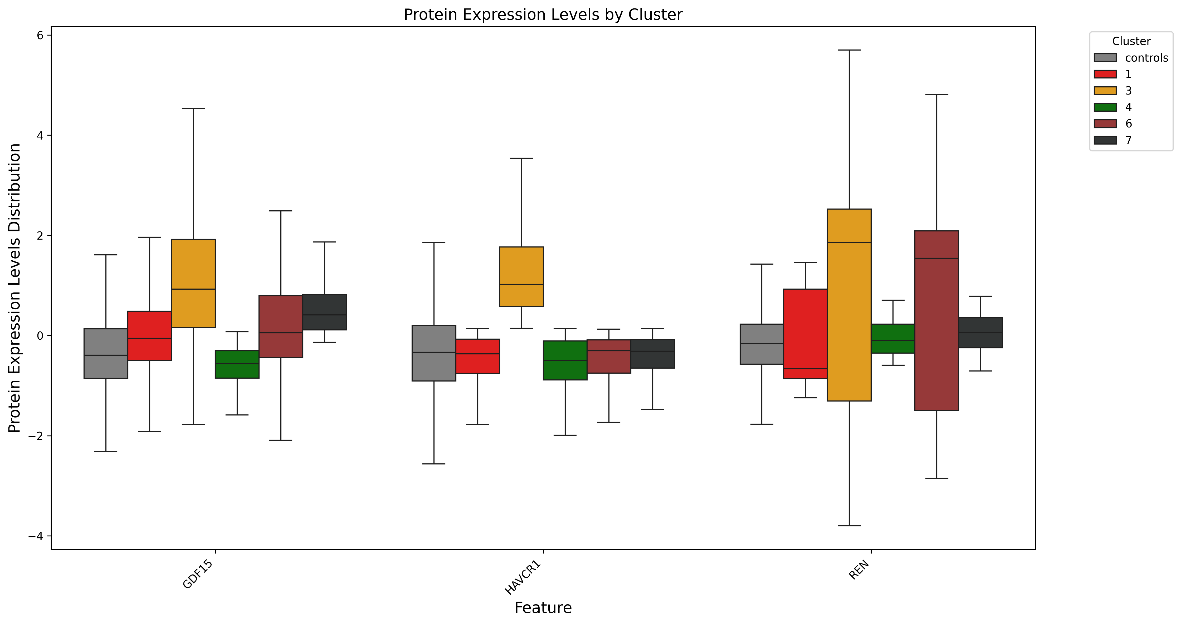

**Figure S4**. Differential expression of separating proteins among hypertension cases.

**Table S17.** Cohen's d effect sizes from Mann-Whitney U tests comparing protein expression levels between cases in each cluster and the cohort of all cases or controls. Upward-pointing arrows indicate up-regulated protein expression; downward-pointing arrows indicate down-regulated protein expression.

| Cluster | GDF15 | | HAVCR1 | | REN | |
| --- | --- | --- | --- | --- | --- | --- |
|  | vs_controls | vs_others | vs_controls | vs_others | vs_controls | vs_others |
| 1 | ▲0.385 | ▼-0.482 | ▼-0.193 | ▼-1.124 | ▲0.113 | ▼-0.326 |
| 2 | ▲1.092 | ▲0.243 | ▲1.629 | ▲0.762 | ▲0.411 | ▼-0.137 |
| 3 | ▲1.559 | ▲0.750 | ▲1.828 | ▲0.966 | ▲1.641 | ▲1.032 |
| 4 | ▼-0.383 | ▼-1.207 | ▼-0.292 | ▼-1.162 | ▲0.146 | ▼-0.298 |
| 5 | ▲0.976 | ▲0.102 | ▲1.520 | ▲0.642 | ▲0.315 | ▼-0.242 |
| 6 | ▲0.576 | ▼-0.278 | ▼-0.156 | ▼-1.002 | ▲0.812 | ▲0.186 |
| 7 | ▲0.974 | ▲0.063 | ▼-0.105 | ▼-1.003 | ▲0.292 | ▼-0.214 |

*Test*

| 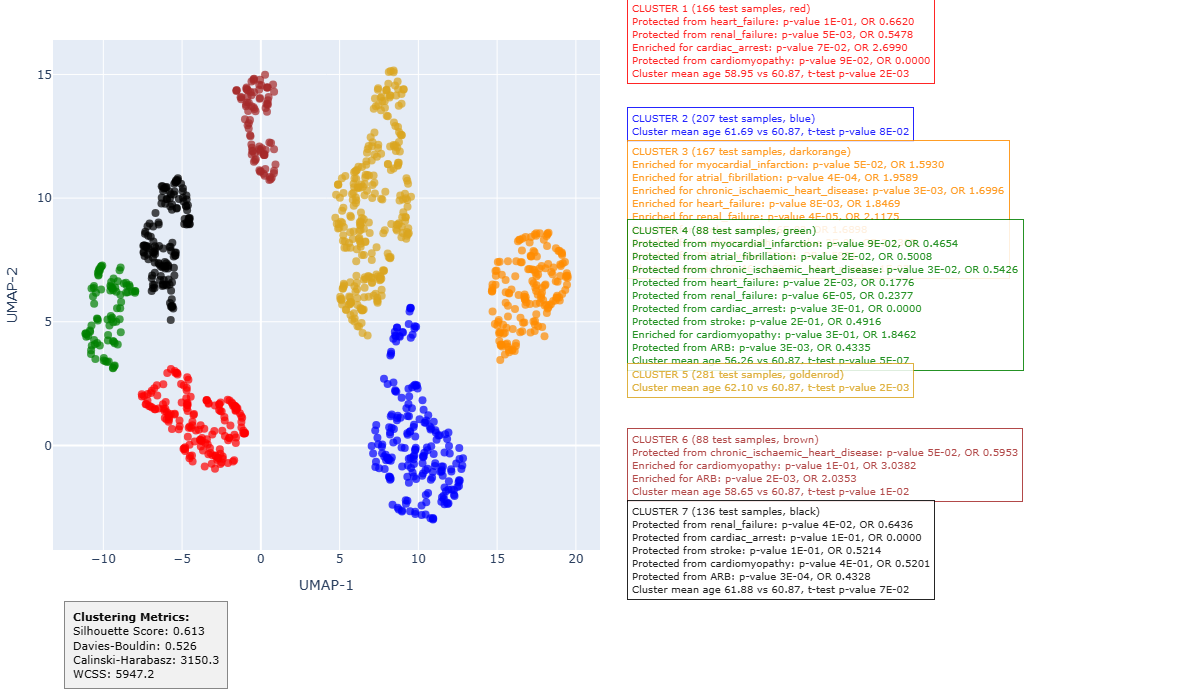 |  |  |
| --- | --- | --- |
|  |  | **CLUSTER 1 (166 test samples, red)** Protected from renal failure: p-value 5E-03, OR 0.5478  Cluster mean age 58.95 vs 60.87, t-test p-value 2E-03 |
|  |  | **CLUSTER 3 (167 test samples, dark orange)** Enriched for atrial fibrillation: p-value 4E-04, OR 1.9589 Enriched for CIHD: p-value 3E-03, OR 1.6996 Enriched for renal failure: p-value 4E-05, OR 2.1175 Cluster mean age 62.46 vs 60.87, t-test p-value 1E-03 |
|  |  | **CLUSTER 4 (88 test samples, green)**  Protected from heart failure: p-value 2E-03, OR 0.1776 Protected from renal failure: p-value 6E-05, OR 0.2377  Cluster mean age 56.26 vs 60.87, t-test p-value 5E-07 |

**Figure S5.** Enrichments for complications, age and sex among hypertension cases from test dataset, fold 2. The silhouette score of the clustering is 0.61. Only complications that were statistically significant in train data according to the criteria in Enrichment analysis, and significant at the *p* < 0.05 level after Benjamini-Hochberg FDR correction are reported.

**Table S18.** Wald test *p*-values and odds ratios for the logistic regression coefficient of each cluster as a predictor of each complication reported in Figure S5 on its own and in the presence of age and sex. The *p*-values in the label, sex, and age model marked bold are significant at the *p* < 0.05 level after Benjamini-Hochberg FDR correction.

| **Cluster** | **Complication** | ***p*-value**  **(label only)** | **Odds ratio**  **(label only)** | ***p*-value**  **(label + sex + age)** | **Odds ratio**  **(label + sex + age)** |
| --- | --- | --- | --- | --- | --- |
| **1** | **renal failure** | **8E-03** | **0.56** | **3E-02** | **0.62** |
| **3** | **atrial fibrillation** | **1E-04** | **1.92** | **2E-03** | **1.69** |
| **3** | **chronic ischaemic heart disease** | **2E-03** | **1.67** | **2E-02** | **1.48** |
| **3** | **renal failure** | **8E-06** | **2.07** | **1E-04** | **1.9** |
| 4 | heart failure | 3E-02 | 0.27 | 7E-02 | 0.35 |
| **4** | **renal failure** | **2E-03** | **0.3** | **1E-02** | **0.36** |

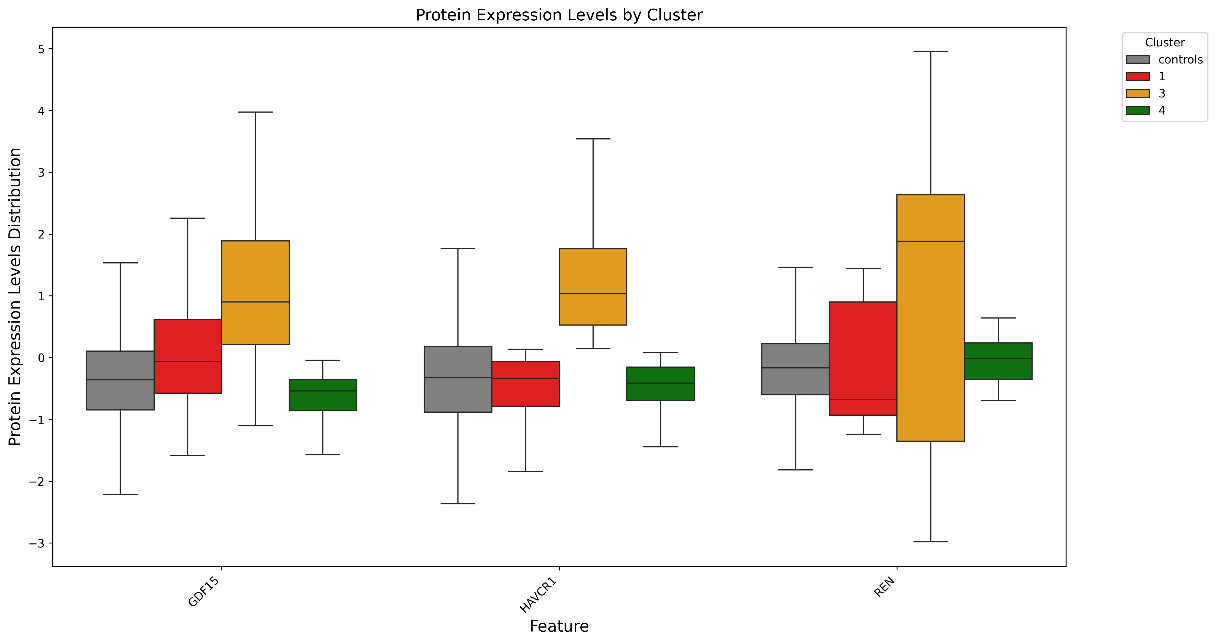

**Figure S6.** Differential expression of separating proteins among hypertension cases.

**Table S19.** Cohen's d effect sizes from Mann-Whitney U tests comparing protein expression levels between cases in each cluster and the cohort of all cases or controls. Upward-pointing arrows indicate up-regulated protein expression; downward-pointing arrows indicate down-regulated protein expression.

| Cluster | GDF15 | | HAVCR1 | | REN | |
| --- | --- | --- | --- | --- | --- | --- |
|  | vs_controls | vs_others | vs_controls | vs_others | vs_controls | vs_others |
| 1 | ▲0.436 | ▼-0.480 | ▼-0.160 | ▼-1.129 | ▲0.080 | ▼-0.374 |
| 2 | ▲1.227 | ▲0.240 | ▲1.755 | ▲0.846 | ▲0.724 | ▲0.055 |
| 3 | ▲1.740 | ▲0.725 | ▲1.952 | ▲1.078 | ▲1.687 | ▲0.994 |
| 4 | ▼-0.404 | ▼-1.135 | ▼-0.183 | ▼-1.038 | ▲0.207 | ▼-0.279 |
| 5 | ▲1.055 | ▲0.068 | ▲1.493 | ▲0.584 | ▲0.347 | ▼-0.254 |
| 6 | ▲0.638 | ▼-0.283 | ▼-0.061 | ▼-0.915 | ▲0.708 | ▲0.080 |
| 7 | ▲1.241 | ▲0.174 | ▼-0.031 | ▼-0.946 | ▲0.344 | ▼-0.212 |

**Fold 3**

*Train*

| 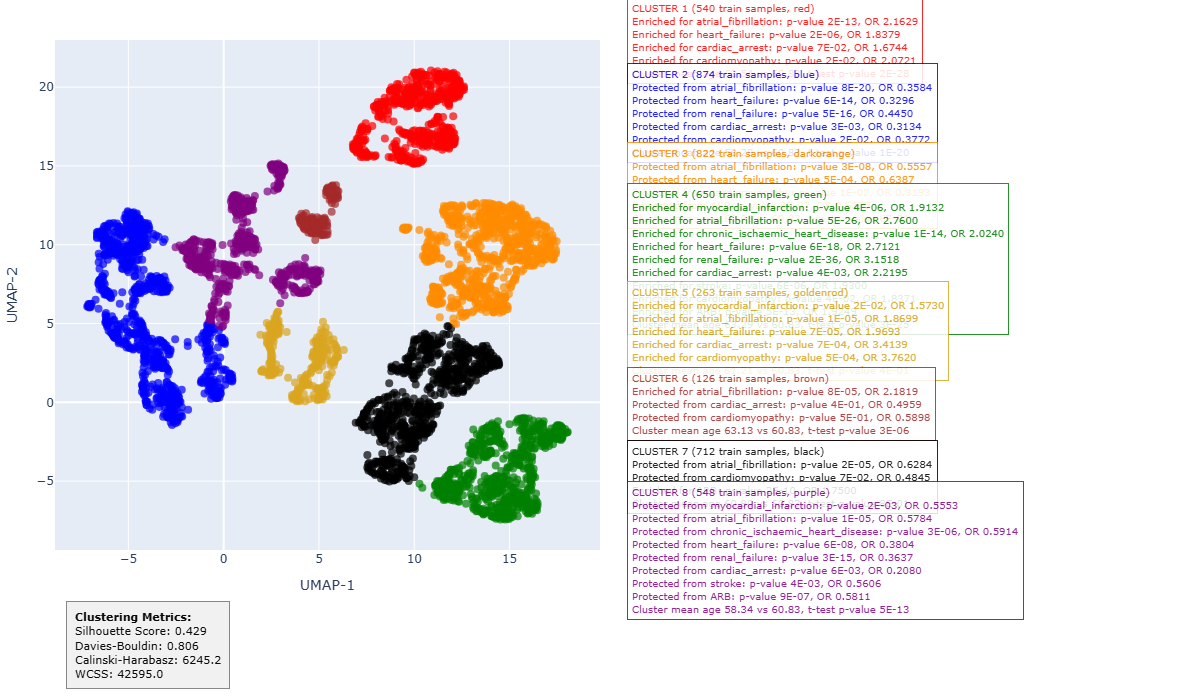 | **CLUSTER 1 (540 train samples, red)** Enriched for atrial fibrillation: p-value 2E-13, OR 2.1629 Enriched for heart failure: p-value 2E-06, OR 1.8379 Enriched for cardiomyopathy: p-value 2E-02, OR 2.0721 Cluster mean age 63.54 vs 60.83, t-test p-value 2E-28 |
| --- | --- |
|  | **CLUSTER 2 (874 train samples, blue)** Protected from atrial fibrillation: p-value 8E-20, OR 0.3584 Protected from heart failure: p-value 6E-14, OR 0.3296 Protected from renal failure: p-value 5E-16, OR 0.4450 Protected from cardiac arrest: p-value 3E-03, OR 0.3134 Protected from cardiomyopathy: p-value 2E-02, OR 0.3772 Cluster mean age 58.29 vs 60.83, t-test p-value 1E-20 |
|  | **CLUSTER 3 (822 train samples, dark orange)** Protected from atrial fibrillation: p-value 3E-08, OR 0.5557 Protected from heart failure: p-value 5E-04, OR 0.6387 Protected from cardiomyopathy: p-value 1E-02, OR 0.3193 |
|  | **CLUSTER 4 (650 train samples, green)** Enriched for myocardial infarction: p-value 4E-06, OR 1.9132 Enriched for atrial fibrillation: p-value 5E-26, OR 2.7600 Enriched for CIHD: p-value 1E-14, OR 2.0240 Enriched for heart failure: p-value 6E-18, OR 2.7121 Enriched for renal failure: p-value 2E-36, OR 3.1518 Enriched for cardiac arrest: p-value 4E-03, OR 2.2195 Enriched for stroke: p-value 6E-06, OR 1.9300 Cluster mean age 63.29 vs 60.83, t-test p-value 3E-25 |
|  | **CLUSTER 5 (263 train samples, goldenrod)** Enriched for myocardial infarction: p-value 2E-02, OR 1.5730 Enriched for atrial fibrillation: p-value 1E-05, OR 1.8699 Enriched for heart failure: p-value 7E-05, OR 1.9693 Enriched for cardiac arrest: p-value 7E-04, OR 3.4139 Enriched for cardiomyopathy: p-value 5E-04, OR 3.7620 |
|  | **CLUSTER 6 (126 train samples, brown)** Enriched for atrial fibrillation: p-value 8E-05, OR 2.1819 Cluster mean age 63.13 vs 60.83, t-test p-value 3E-06 |
|  | **CLUSTER 7 (712 train samples, black)** Protected from atrial fibrillation: p-value 2E-05, OR 0.6284 |
|  | **CLUSTER 8 (548 train samples, purple)**  Protected from myocardial infarction: p-value 2E-03, OR 0.5553  Protected from atrial fibrillation: p-value 1E-05, OR 0.5784  Protected from CIHD: p-value 3E-06, OR 0.5914  Protected from heart failure: p-value 6E-08, OR 0.3804  Protected from renal failure: p-value 3E-15, OR 0.3637  Protected from cardiac arrest: p-value 6E-03, OR 0.2080  Protected from stroke: p-value 4E-03, OR 0.5606  Cluster mean age 58.34 vs 60.83, t-test p-value 5E-13 |

**Figure S7.** Enrichments for complications, age and sex among hypertension cases from training dataset, fold 3. The silhouette score of the clustering is 0.43.

**Table S20.** Wald test *p*-values and odds ratios for the logistic regression coefficient of each cluster as a predictor of each complication reported in Figure S7 on its own and in the presence of age and sex. The *p*-values in the label, sex, and age model marked bold are significant at the *p* < 0.05 level after Benjamini-Hochberg FDR correction.

| **Cluster** | **Complication** | ***p*-value**  **(label only)** | **Odds ratio**  **(label only)** | ***p*-value**  **(label+sex+age)** | **Odds ratio**  **(label+sex+age)** |
| --- | --- | --- | --- | --- | --- |
| **1** | **atrial fibrillation** | **2E-16** | **2.15** | **3E-11** | **1.88** |
| **1** | **heart failure** | **1E-07** | **1.82** | **6E-06** | **1.67** |
| **1** | **cardiomyopathy** | **2E-02** | **1.94** | **8E-03** | **2.17** |
| **2** | **atrial fibrillation** | **0E+00** | **0.36** | **2E-13** | **0.41** |
| **2** | **heart failure** | **2E-11** | **0.34** | **9E-10** | **0.37** |
| **2** | **renal failure** | **2E-15** | **0.45** | **7E-11** | **0.52** |
| **2** | **cardiac arrest** | **2E-02** | **0.38** | **4E-02** | **0.42** |
| **2** | **cardiomyopathy** | **5E-02** | **0.45** | **3E-02** | **0.41** |
| **3** | **atrial fibrillation** | **3E-08** | **0.56** | **1E-08** | **0.55** |
| **3** | **heart failure** | **6E-04** | **0.64** | **5E-04** | **0.64** |
| **3** | **cardiomyopathy** | **4E-02** | **0.4** | **4E-02** | **0.4** |
| **4** | **myocardial infarction** | **1E-07** | **1.89** | **2E-06** | **1.78** |
| **4** | **atrial fibrillation** | **0E+00** | **2.75** | **0E+00** | **2.36** |
| **4** | **chronic ischaemic heart disease** | **0E+00** | **2.02** | **2E-13** | **1.85** |
| **4** | **heart failure** | **0E+00** | **2.68** | **0E+00** | **2.45** |
| **4** | **renal failure** | **0E+00** | **3.13** | **0E+00** | **2.79** |
| **4** | **cardiac arrest** | **2E-03** | **2.1** | **7E-03** | **1.88** |
| **4** | **stroke** | **2E-07** | **1.91** | **7E-06** | **1.75** |
| **5** | **myocardial infarction** | **3E-02** | **1.54** | **2E-02** | **1.57** |
| **5** | **atrial fibrillation** | **3E-06** | **1.88** | **3E-06** | **1.92** |
| **5** | **heart failure** | **3E-05** | **1.93** | **3E-05** | **1.94** |
| **5** | **cardiac arrest** | **4E-04** | **3.01** | **4E-04** | **3** |
| **5** | **cardiomyopathy** | **3E-04** | **3.25** | **2E-04** | **3.33** |
| **6** | **atrial fibrillation** | **7E-05** | **2.13** | **1E-03** | **1.86** |
| **7** | **atrial fibrillation** | **2E-05** | **0.63** | **6E-06** | **0.61** |
| **8** | **myocardial infarction** | **8E-03** | **0.59** | **3E-02** | **0.66** |
| **8** | **atrial fibrillation** | **4E-05** | **0.59** | **1E-02** | **0.73** |
| **8** | **chronic ischaemic heart disease** | **6E-06** | **0.6** | **2E-03** | **0.7** |
| **8** | **heart failure** | **1E-06** | **0.39** | **3E-05** | **0.44** |
| **8** | **renal failure** | **4E-13** | **0.37** | **4E-10** | **0.42** |
| 8 | cardiac arrest | 5E-02 | 0.33 | 8E-02 | 0.37 |
| **8** | **stroke** | **8E-03** | **0.57** | **3E-02** | **0.64** |

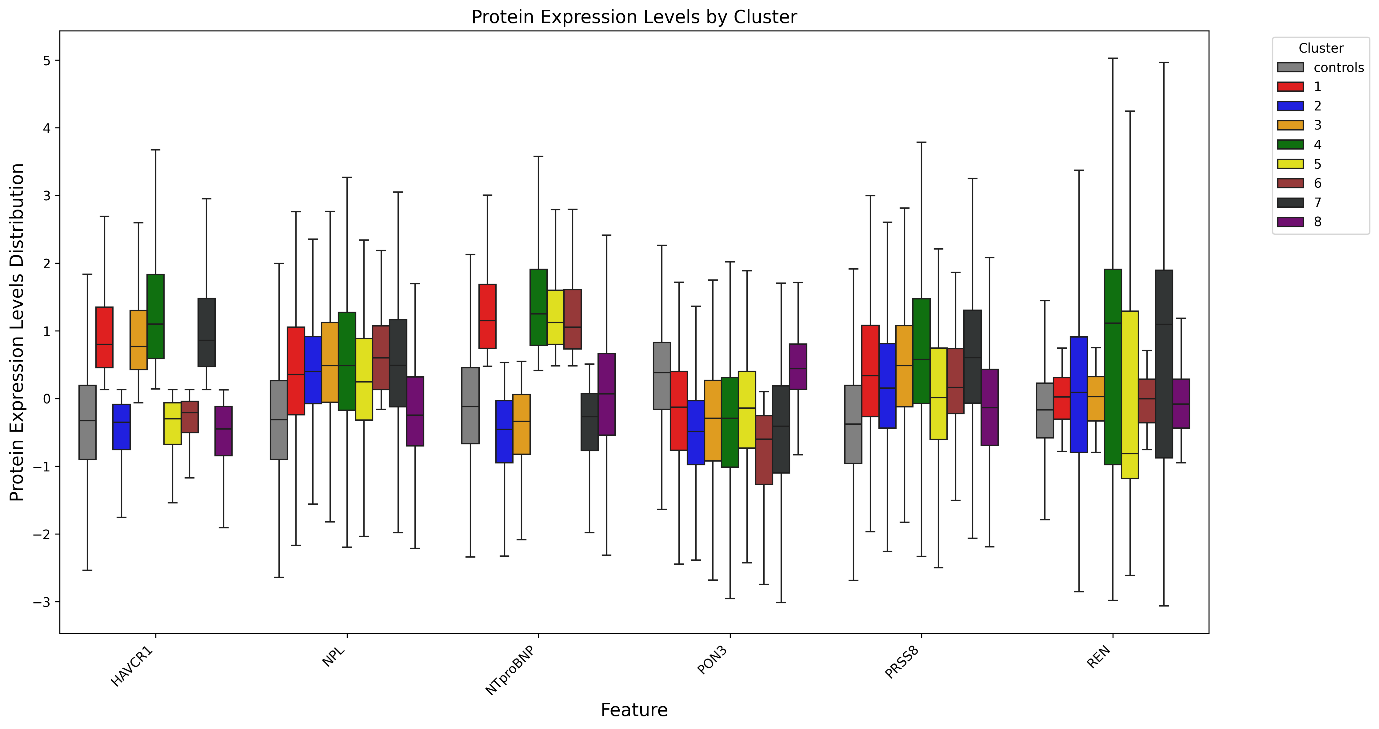

**Figure S8.** Differential expression of separating proteins among hypertension cases.

**Table S21.** Cohen's d effect sizes from Mann-Whitney U tests comparing protein expression levels between cases in each cluster and the cohort of all cases or controls. Upward-pointing arrows in red indicate up-regulated protein expression; downward-pointing arrows in blue indicate down-regulated protein expression.

| Cluster | HAVCR1 | | NPL | | NTproBNP | | PON3 | | PRSS8 | | REN | |
| --- | --- | --- | --- | --- | --- | --- | --- | --- | --- | --- | --- | --- |
|  | vs_controls | vs_others | vs_controls | vs_others | vs_controls | vs_others | vs_controls | vs_others | vs_controls | vs_others | vs_controls | vs_others |
| 1 | *▲*1.538 | *▲*0.581 | *▲*0.777 | *▲*0.030 | *▲*1.672 | *▲*1.147 | ▼-0.608 | ▲0.086 | ▲0.869 | ▲0.074 | ▲0.227 | ▼-0.273 |
| 2 | *▼*-0.188 | *▼*-1.237 | *▲*0.748 | *▼*-0.007 | *▼*-0.510 | *▼*-0.931 | ▼-0.984 | ▼-0.293 | ▲0.592 | ▼-0.221 | ▲0.480 | ▼-0.082 |
| 3 | *▲*1.493 | *▲*0.571 | *▲*0.894 | *▲*0.167 | *▼*-0.388 | *▼*-0.782 | ▼-0.753 | ▼-0.076 | ▲0.923 | ▲0.135 | ▲0.235 | ▼-0.286 |
| 4 | *▲*1.904 | *▲*1.036 | *▲*0.896 | *▲*0.167 | *▲*1.839 | *▲*1.422 | ▼-0.774 | ▼-0.081 | ▲1.128 | ▲0.356 | ▲1.161 | ▲0.477 |
| 5 | *▼*-0.096 | *▼*-0.907 | *▲*0.655 | *▼*-0.097 | *▲*1.635 | *▲*0.999 | ▼-0.540 | ▲0.145 | ▲0.430 | ▼-0.344 | ▲0.325 | ▼-0.180 |
| 6 | *▲*0.029 | *▼*-0.762 | *▲*1.088 | *▲*0.320 | *▲*1.524 | *▲*0.864 | ▼-1.258 | ▼-0.466 | ▲0.696 | ▼-0.097 | ▲0.198 | ▼-0.260 |
| 7 | *▲*1.606 | *▲*0.685 | *▲*0.891 | *▲*0.164 | *▼*-0.345 | *▼*-0.727 | ▼-0.853 | ▼-0.159 | ▲1.075 | ▲0.295 | ▲1.267 | ▲0.560 |
| 8 | *▼*-0.287 | *▼*-1.208 | *▲*0.140 | *▼*-0.669 | *▲*0.227 | *▼*-0.177 | ▲0.112 | ▲0.791 | ▲0.280 | ▼-0.525 | ▲0.147 | ▼-0.322 |

*Test*

| 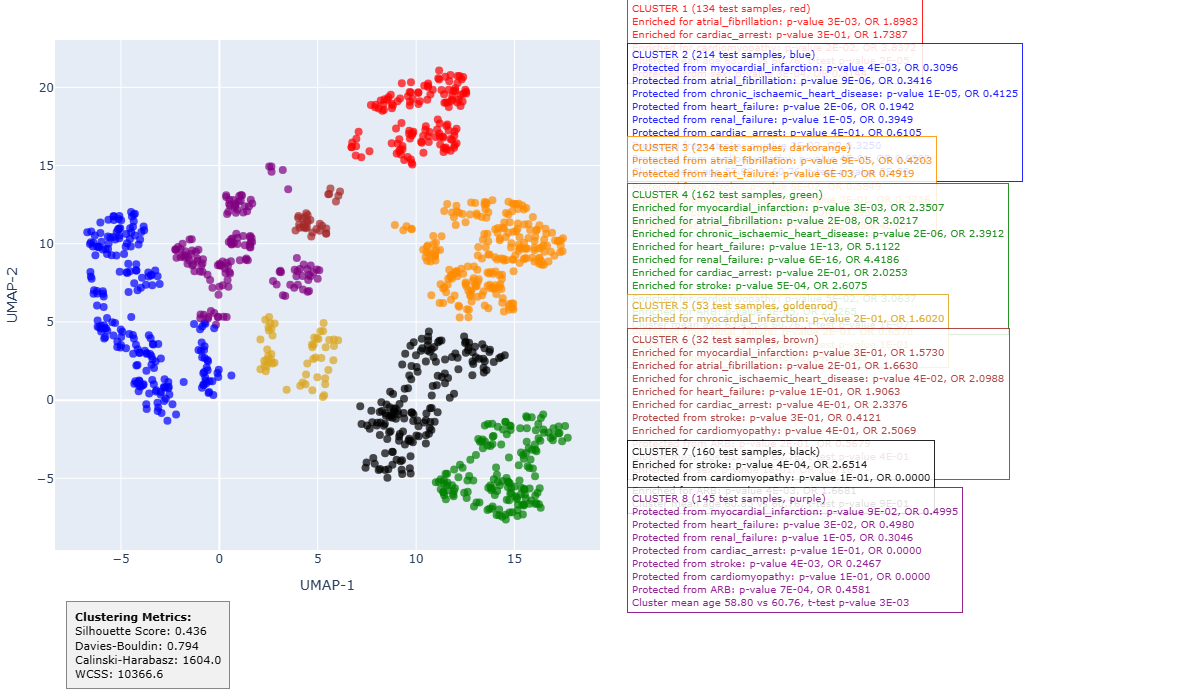 | **CLUSTER 1 (134 test samples, red)** Enriched for atrial fibrillation: p-value 3E-03, OR 1.8983  Cluster mean age 63.07 vs 60.76, t-test p-value 2E-05 Protected from sex: p-value 3E-04, OR 0.5144 |
| --- | --- |
|  | **CLUSTER 2 (214 test samples, blue)**  Protected from atrial fibrillation: p-value 9E-06, OR 0.3416 Protected from heart failure: p-value 2E-06, OR 0.1942 Protected from renal failure: p-value 1E-05, OR 0.3949  Cluster mean age 57.69 vs 60.76, t-test p-value 3E-08 |
|  | **CLUSTER 3 (234 test samples, dark orange)** Protected from atrial fibrillation: p-value 9E-05, OR 0.4203 Protected from heart failure: p-value 6E-03, OR 0.4919 |
|  | **CLUSTER 4 (162 test samples, green)** Enriched for myocardial infarction: p-value 3E-03, OR 2.3507 Enriched for atrial fibrillation: p-value 2E-08, OR 3.0217 Enriched for CIHD: p-value 2E-06, OR 2.3912 Enriched for heart failure: p-value 1E-13, OR 5.1122 Enriched for renal failure: p-value 6E-16, OR 4.4186  Enriched for stroke: p-value 5E-04, OR 2.6075  Cluster mean age 63.43 vs 60.76, t-test p-value 1E-07 |
|  | **CLUSTER 8 (145 test samples, purple)**  Protected from renal failure: p-value 1E-05, OR 0.3046 Protected from stroke: p-value 4E-03, OR 0.2467  Cluster mean age 58.80 vs 60.76, t-test p-value 3E-03 |

**Figure S9.** Enrichments for complications, age and sex among hypertension cases from test dataset, fold 3. The silhouette score of the clustering is 0.44. Only complications that were statistically significant in train data according to the criteria in Enrichment analysis, and significant at the *p* < 0.05 level after Benjamini-Hochberg FDR correction are reported.

**Table S22.** Wald test *p*-values and odds ratios for the logistic regression coefficient of each cluster as a predictor of each complication reported in Figure S9 on its own and in the presence of age and sex. The *p*-values in the label, sex, and age model marked bold are significant at the *p* < 0.05 level after Benjamini-Hochberg FDR correction.

| **Cluster** | **Complication** | ***p*-value**  **(label only)** | **Odds ratio**  **(label only)** | ***p*-value**  **(label+sex+age)** | **Odds ratio**  **(label+sex+age)** |
| --- | --- | --- | --- | --- | --- |
| **1** | **atrial fibrillation** | **2E-03** | **1.84** | **1E-02** | **1.68** |
| **2** | **atrial fibrillation** | **7E-05** | **0.37** | **2E-03** | **0.46** |
| **2** | **heart failure** | **2E-04** | **0.24** | **1E-03** | **0.30** |
| **2** | **renal failure** | **5E-05** | **0.41** | **5E-04** | **0.46** |
| **3** | **atrial fibrillation** | **3E-04** | **0.44** | **9E-05** | **0.41** |
| **3** | **heart failure** | **1E-02** | **0.52** | **5E-03** | **0.48** |
| **4** | **myocardial infarction** | **1E-03** | **2.20** | **3E-03** | **2.10** |
| **4** | **atrial fibrillation** | **2E-10** | **2.91** | **2E-07** | **2.42** |
| **4** | **chronic ischaemic heart disease** | **2E-07** | **2.33** | **4E-06** | **2.17** |
| **4** | **heart failure** | **0E+00** | **4.78** | **2E-15** | **4.13** |
| **4** | **renal failure** | **0E+00** | **4.22** | **0E+00** | **3.92** |
| **4** | **stroke** | **9E-05** | **2.45** | **7E-04** | **2.18** |
| **8** | **renal failure** | **2E-04** | **0.34** | **7E-04** | **0.37** |
| 8 | stroke | 3E-02 | 0.34 | 6E-02 | 0.38 |

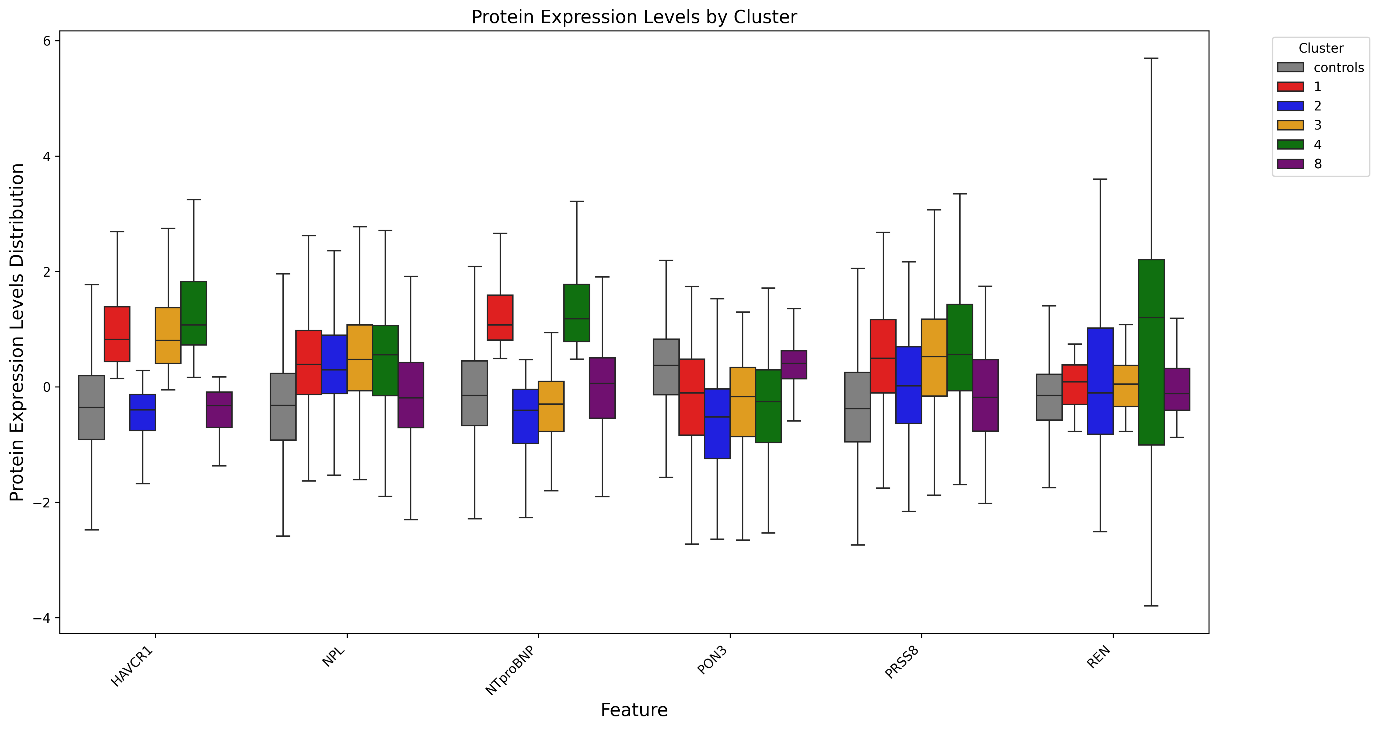
**Figure S10.** Differential expression of separating proteins among hypertension cases.

**Table S23.** Cohen's d effect sizes from Mann-Whitney U tests comparing protein expression levels between cases in each cluster and the cohort of all cases or controls. Upward-pointing arrows indicate up-regulated protein expression; downward-pointing arrows indicate down-regulated protein expression.

| Cluster | HAVCR1 | | NPL | | NTproBNP | | PON3 | | PRSS8 | | REN | |
| --- | --- | --- | --- | --- | --- | --- | --- | --- | --- | --- | --- | --- |
|  | vs_controls | vs_others | vs_controls | vs_others | vs_controls | vs_others | vs_controls | vs_others | vs_controls | vs_others | vs_controls | vs_others |
| 1 | ▲1.615 | ▲0.631 | ▲0.829 | ▲0.087 | ▲1.714 | ▲1.167 | ▼-0.495 | ▲0.136 | ▲0.978 | ▲0.211 | ▲0.266 | ▼-0.201 |
| 2 | ▼-0.179 | ▼-1.324 | ▲0.745 | ▼-0.008 | ▼-0.479 | ▼-0.872 | ▼-0.952 | ▼-0.399 | ▲0.395 | ▼-0.399 | ▲0.338 | ▼-0.139 |
| 3 | ▲1.536 | ▲0.593 | ▲0.864 | ▲0.139 | ▼-0.348 | ▼-0.751 | ▼-0.658 | ▼-0.021 | ▲0.939 | ▲0.186 | ▲0.261 | ▼-0.226 |
| 4 | ▲1.98 | ▲1.091 | ▲0.924 | ▲0.210 | ▲1.859 | ▲1.438 | ▼-0.712 | ▼-0.073 | ▲1.088 | ▲0.338 | ▲1.188 | ▲0.570 |
| 5 | ▲0.008 | ▼-0.868 | ▲0.238 | ▼-0.516 | ▲1.647 | ▲0.973 | ▼-0.461 | ▲0.153 | ▲0.408 | ▼-0.329 | ▼-0.249 | ▼-0.493 |
| 6 | ▼-0.138 | ▼-0.977 | ▲1.096 | ▲0.339 | ▲1.809 | ▲1.083 | ▼-1.138 | ▼-0.402 | ▲0.895 | ▲0.111 | ▲0.105 | ▼-0.272 |
| 7 | ▲1.566 | ▲0.583 | ▲0.944 | ▲0.229 | ▼-0.366 | ▼-0.713 | ▼-0.790 | ▼-0.142 | ▲1.133 | ▲0.390 | ▲1.217 | ▲0.555 |
| 8 | ▼-0.112 | ▼-1.117 | ▲0.211 | ▼-0.613 | ▲0.158 | ▼-0.233 | ▲0.115 | ▲0.713 | ▲0.158 | ▼-0.620 | ▲0.174 | ▼-0.260 |

**Fold 4**

*Train*

| 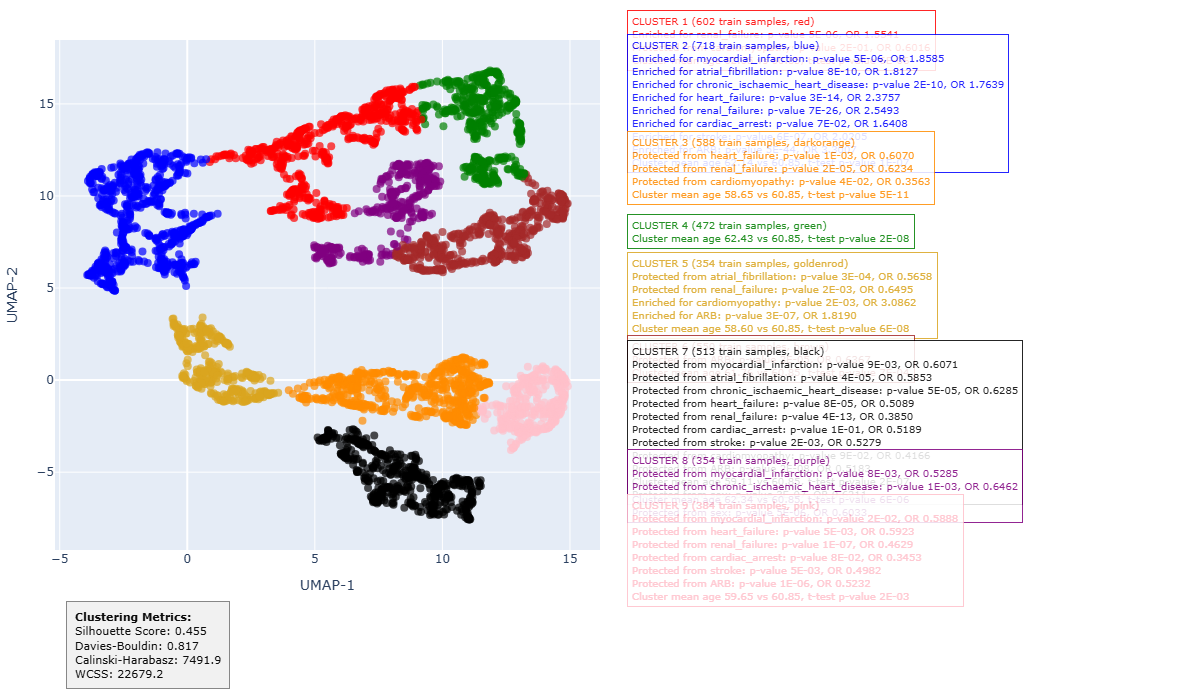 | **CLUSTER 1 (602 train samples, red)** Enriched for renal failure: p-value 5E-06, OR 1.5541 Cluster mean age 62.05 vs 60.85, t-test p-value 6E-06 |
| --- | --- |
|  | **CLUSTER 2 (718 train samples, blue)** Enriched for myocardial infarction: p-value 5E-06, OR 1.8585 Enriched for atrial fibrillation: p-value 8E-10, OR 1.8127 Enriched for CIHD: p-value 2E-10, OR 1.7639 Enriched for heart failure: p-value 3E-14, OR 2.3757 Enriched for renal failure: p-value 7E-26, OR 2.5493 Enriched for stroke: p-value 6E-07, OR 2.0205 Cluster mean age 62.14 vs 60.85, t-test p-value 1E-07 |
|  | **CLUSTER 3 (588 train samples, dark orange)** Protected from heart failure: p-value 1E-03, OR 0.6070 Protected from renal failure: p-value 2E-05, OR 0.6234  Cluster mean age 58.65 vs 60.85, t-test p-value 5E-11 |
|  | **CLUSTER 5 (354 train samples, goldenrod)** Protected from atrial fibrillation: p-value 3E-04, OR 0.5658 Protected from renal failure: p-value 2E-03, OR 0.6495 Enriched for cardiomyopathy: p-value 2E-03, OR 3.0862 Cluster mean age 58.60 vs 60.85, t-test p-value 6E-08 |
|  | **CLUSTER 7 (513 train samples, black)** Protected from myocardial infarction: p-value 9E-03, OR 0.6071 Protected from atrial fibrillation: p-value 4E-05, OR 0.5853 Protected from CIHD: p-value 5E-05, OR 0.6285 Protected from heart failure: p-value 8E-05, OR 0.5089 Protected from renal failure: p-value 4E-13, OR 0.3850 Protected from stroke: p-value 2E-03, OR 0.5279 Cluster mean age 59.11 vs 60.85, t-test p-value 2E-07 Protected from sex: p-value 3E-07, OR 0.6211 |
|  | **CLUSTER 8 (354 train samples, purple)** Protected from myocardial infarction: p-value 8E-03, OR 0.5285 Protected from CIHD: p-value 1E-03, OR 0.6462 Cluster mean age 62.34 vs 60.85, t-test p-value 6E-06 Protected from sex: p-value 5E-06, OR 0.6033 |
|  | **CLUSTER 9 (384 train samples, pink)**  Protected from myocardial infarction: p-value 2E-02, OR 0.5888  Protected from heart failure: p-value 5E-03, OR 0.5923  Protected from renal failure: p-value 1E-07, OR 0.4629  Protected from stroke: p-value 5E-03, OR 0.4982  Cluster mean age 59.65 vs 60.85, t-test p-value 2E-03 |

**Figure S11.** Enrichments for complications, age and sex among hypertension cases from training dataset, fold 4. The silhouette score of the clustering is 0.46.

**Table S24.** Wald test *p*-values and odds ratios for the logistic regression coefficient of each cluster as a predictor of each complication reported in Figure S11 on its own and in the presence of age and sex. The *p*-values in the label, sex, and age model marked bold are significant at the *p* < 0.05 level after Benjamini-Hochberg FDR correction.

| **Cluster** | **Complication** | ***p*-value**  **(label only)** | **Odds ratio**  **(label only)** | ***p*-value**  **(label + sex + age)** | **Odds ratio**  **(label + sex + age)** |
| --- | --- | --- | --- | --- | --- |
| **1** | **renal failure** | **4E-07** | **1.57** | **1E-05** | **1.48** |
| **2** | **myocardial infarction** | **2E-07** | **1.84** | **5E-06** | **1.71** |
| **2** | **atrial fibrillation** | **2E-12** | **1.81** | **5E-08** | **1.60** |
| **2** | **chronic ischaemic heart disease** | **8E-13** | **1.76** | **8E-09** | **1.59** |
| **2** | **heart failure** | **0E+00** | **2.35** | **7E-16** | **2.16** |
| **2** | **renal failure** | **0E+00** | **2.54** | **0E+00** | **2.37** |
| **2** | **stroke** | **5E-09** | **2.00** | **2E-07** | **1.85** |
| **3** | **heart failure** | **2E-03** | **0.62** | **2E-02** | **0.68** |
| **3** | **renal failure** | **4E-05** | **0.63** | **2E-03** | **0.70** |
| **5** | **atrial fibrillation** | **7E-04** | **0.57** | **2E-02** | **0.67** |
| **5** | **renal failure** | **3E-03** | **0.65** | **4E-02** | **0.74** |
| **5** | **cardiomyopathy** | **1E-03** | **2.75** | **1E-03** | **2.66** |
| 7 | myocardial infarction | 2E-02 | 0.62 | 8E-02 | 0.70 |
| **7** | **atrial fibrillation** | **1E-04** | **0.59** | **1E-02** | **0.71** |
| **7** | **chronic ischaemic heart disease** | **9E-05** | **0.63** | **1E-02** | **0.74** |
| **7** | **heart failure** | **3E-04** | **0.52** | **4E-03** | **0.60** |
| **7** | **renal failure** | **3E-11** | **0.39** | **4E-09** | **0.44** |
| **7** | **stroke** | **7E-03** | **0.55** | **3E-02** | **0.61** |
| **8** | **myocardial infarction** | **2E-02** | **0.56** | **3E-02** | **0.58** |
| **8** | **chronic ischaemic heart disease** | **8E-04** | **0.62** | **1E-03** | **0.63** |
| **9** | **myocardial infarction** | **3E-02** | **0.60** | **3E-02** | **0.60** |
| **9** | **heart failure** | **1E-02** | **0.60** | **2E-02** | **0.62** |
| **9** | **renal failure** | **7E-07** | **0.47** | **3E-06** | **0.49** |
| **9** | **stroke** | **1E-02** | **0.52** | **2E-02** | **0.54** |

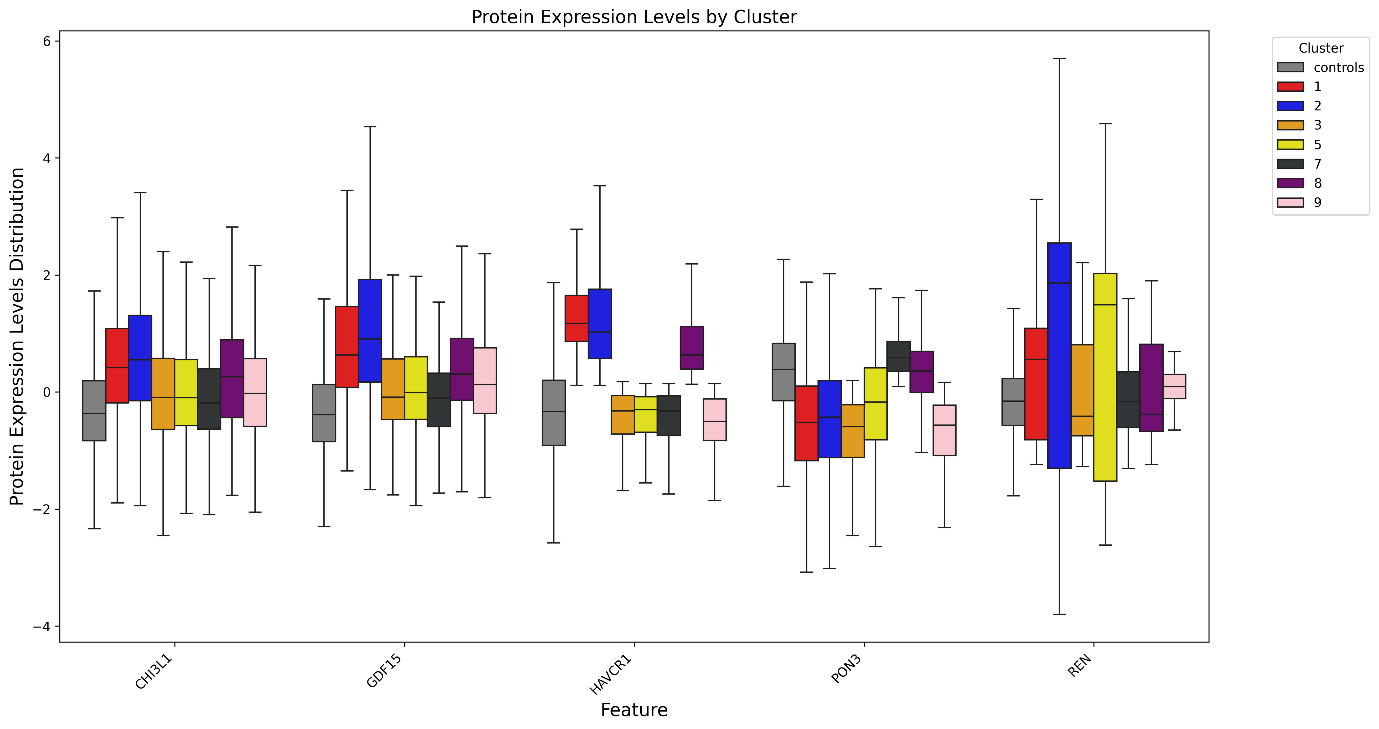
**Figure S12.** Differential expression of separating proteins among hypertension cases.

**Table S25.** Cohen's d effect sizes from Mann-Whitney U tests comparing protein expression levels between cases in each cluster and the cohort of all cases or controls. Upward-pointing arrows indicate up-regulated protein expression; downward-pointing arrows indicate down-regulated protein expression.

| Cluster | CHI3L1 | | GDF15 | | HAVCR1 | | PON3 | | REN | |
| --- | --- | --- | --- | --- | --- | --- | --- | --- | --- | --- |
|  | vs_controls | vs_others | vs_controls | vs_others | vs_controls | vs_others | vs_controls | vs_others | vs_controls | vs_others |
| 1 | ▲0.916 | ▲0.240 | ▲1.335 | ▲0.401 | ▲1.945 | ▲1.018 | ▼-0.954 | ▼-0.255 | ▲0.441 | ▼-0.107 |
| 2 | ▲1.041 | ▲0.373 | ▲1.606 | ▲0.727 | ▲1.849 | ▲0.986 | ▼-0.872 | ▼-0.182 | ▲1.656 | ▲1.044 |
| 3 | ▲0.356 | ▼-0.292 | ▲0.433 | ▼-0.449 | ▼-0.148 | ▼-1.075 | ▼-1.197 | ▼-0.466 | ▲0.175 | ▼-0.284 |
| 4 | ▲1.077 | ▲0.373 | ▲1.409 | ▲0.435 | ▲2.079 | ▲1.105 | ▼-0.808 | ▼-0.104 | ▲0.374 | ▼-0.162 |
| 5 | ▲0.377 | ▼-0.262 | ▲0.491 | ▼-0.375 | ▼-0.129 | ▼-0.979 | ▼-0.680 | ▼-0.003 | ▲0.652 | ▲0.074 |
| 6 | ▲0.680 | ▲0.008 | ▲0.732 | ▼-0.182 | ▲0.891 | ▼-0.060 | ▼-0.755 | ▼-0.052 | ▲0.404 | ▼-0.145 |
| 7 | ▲0.214 | ▼-0.423 | ▲0.271 | ▼-0.597 | ▼-0.165 | ▼-1.077 | ▲0.442 | ▲1.135 | ▲0.053 | ▼-0.361 |
| 8 | ▲0.700 | ▲0.023 | ▲0.855 | ▼-0.063 | ▲1.391 | ▲0.411 | ▼-0.085 | ▲0.565 | ▲0.241 | ▼-0.230 |
| 9 | ▲0.362 | ▼-0.278 | ▲0.598 | ▼-0.289 | ▼-0.231 | ▼-1.095 | ▼-1.151 | ▼-0.399 | ▲0.364 | ▼-0.163 |

*Test*

| 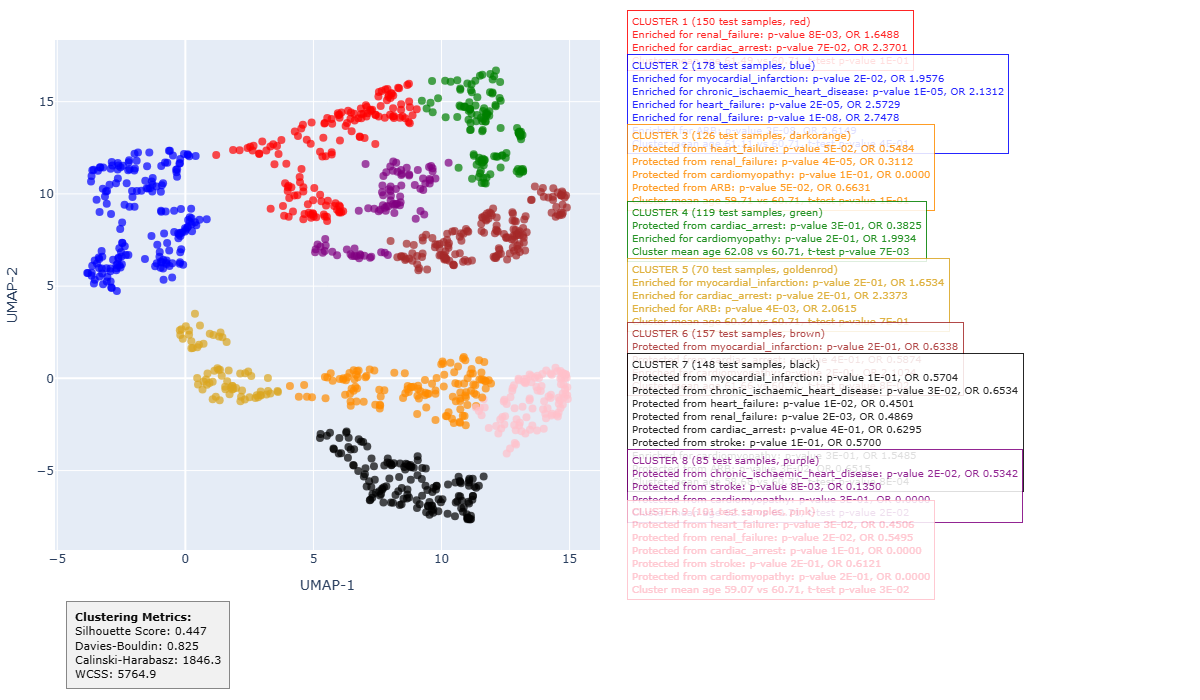 |  |  |
| --- | --- | --- |
|  |  | **CLUSTER 2 (178 test samples, blue)**  Enriched for CIHD: p-value 1E-05, OR 2.1312 Enriched for heart failure: p-value 2E-05, OR 2.5729 Enriched for renal failure: p-value 1E-08, OR 2.7478 |
|  |  | **CLUSTER 3 (126 test samples, dark orange)**  Protected from renal failure: p-value 4E-05, OR 0.3112 |
|  |  | **CLUSTER 7 (148 test samples, black)**  Protected from renal failure: p-value 2E-03, OR 0.4869 Cluster mean age 58.68 vs 60.71, t-test p-value 8E-04 |

**Figure S13.** Enrichments for complications, age and sex among hypertension cases from test dataset, fold 4. The silhouette score of the clustering is 0.45. Only complications that were statistically significant in train data according to the criteria in Enrichment analysis, and significant at the *p* < 0.05 level after Benjamini-Hochberg FDR correction are reported.

**Table S26.** Wald test *p*-values and odds ratios for the logistic regression coefficient of each cluster as a predictor of each complication reported in Figure S13 on its own and in the presence of age and sex. The *p*-values in the label, sex, and age model marked bold are significant at the *p* < 0.05 level after Benjamini-Hochberg FDR correction.

| **Cluster** | **Complication** | ***p*-value**  **(label only)** | **Odds ratio**  **(label only)** | ***p*-value**  **(label+sex+age)** | **Odds ratio**  **(label+sex+age)** |
| --- | --- | --- | --- | --- | --- |
| **2** | **chronic ischaemic heart disease** | **2E-06** | **2.09** | **7E-06** | **2.04** |
| **2** | **heart failure** | **7E-07** | **2.47** | **2E-06** | **2.42** |
| **2** | **renal failure** | **2E-10** | **2.67** | **5E-10** | **2.65** |
| **3** | **renal failure** | **5E-04** | **0.35** | **8E-04** | **0.36** |
| **7** | **renal failure** | **8E-03** | **0.54** | **3E-02** | **0.60** |

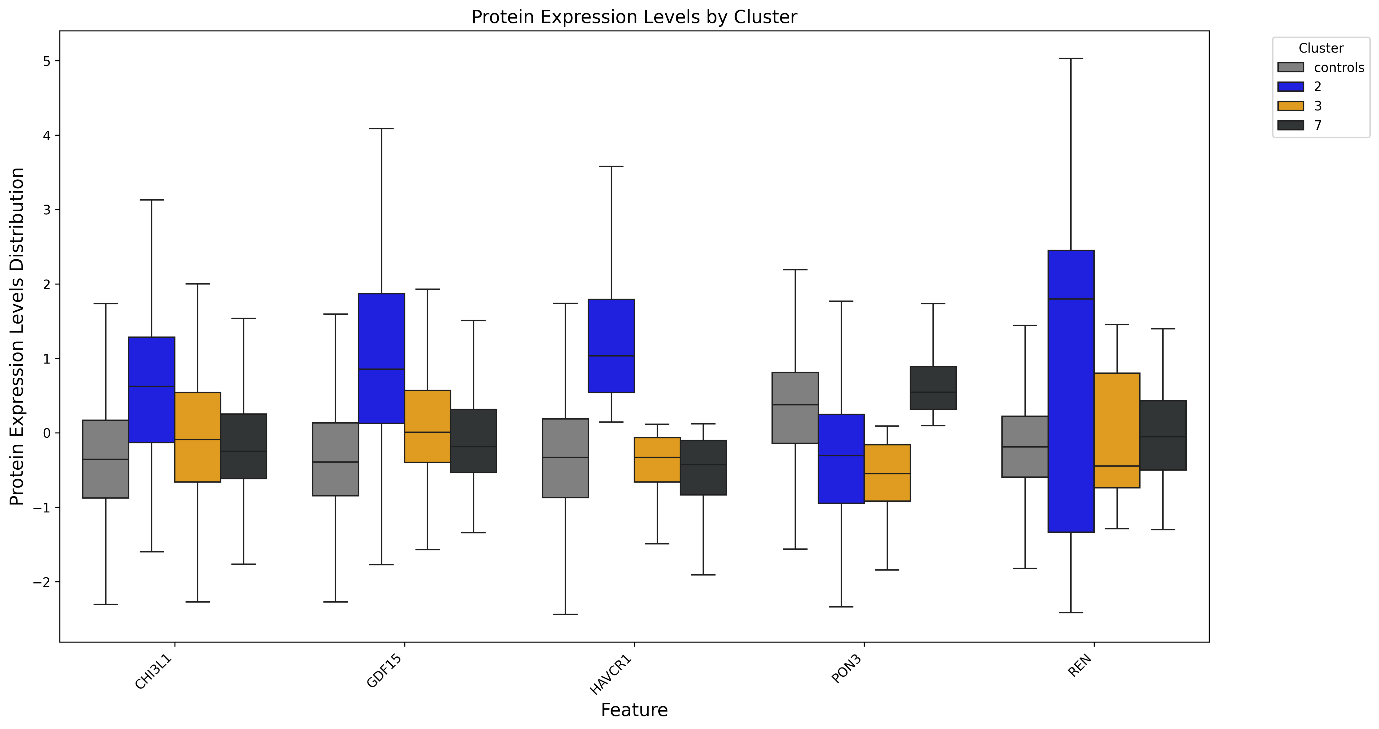
**Figure S14.** Differential expression of separating proteins among hypertension cases.

**Table S27.** Cohen's d effect sizes from Mann-Whitney U tests comparing protein expression levels between cases in each cluster and the cohort of all cases or controls. Upward-pointing arrows indicate up-regulated protein expression; downward-pointing arrows indicate down-regulated protein expression.

| Cluster | CHI3L1 | | GDF15 | | HAVCR1 | | PON3 | | REN | |
| --- | --- | --- | --- | --- | --- | --- | --- | --- | --- | --- |
|  | vs_controls | vs_others | vs_controls | vs_others | vs_controls | vs_others | vs_controls | vs_others | vs_controls | vs_others |
| 1 | ▲0.879 | ▲0.218 | ▲1.172 | ▲0.360 | ▲1.919 | ▲0.987 | ▼-0.806 | ▼-0.154 | ▲0.411 | ▼-0.165 |
| 2 | ▲1.079 | ▲0.421 | ▲1.484 | ▲0.786 | ▲1.844 | ▲0.990 | ▼-0.718 | ▼-0.079 | ▲1.533 | ▲0.883 |
| 3 | ▲0.327 | ▼-0.296 | ▲0.415 | ▼-0.419 | ▼-0.120 | ▼-1.021 | ▼-1.162 | ▼-0.449 | ▲0.213 | ▼-0.289 |
| 4 | ▲0.856 | ▲0.181 | ▲1.117 | ▲0.285 | ▲2.035 | ▲1.059 | ▼-0.784 | ▼-0.190 | ▲0.574 | ▼-0.078 |
| 5 | ▲0.448 | ▼-0.183 | ▲0.826 | ▼-0.003 | ▼-0.121 | ▼-0.952 | ▼-0.713 | ▼-0.060 | ▲1.106 | ▲0.345 |
| 6 | ▲0.684 | ▲0.027 | ▲0.663 | ▼-0.178 | ▲0.907 | ▼-0.054 | ▼-0.799 | ▼-0.144 | ▲0.393 | ▼-0.194 |
| 7 | ▲0.177 | ▼-0.446 | ▲0.189 | ▼-0.675 | ▼-0.278 | ▼-1.240 | ▲0.462 | ▲1.019 | ▲0.207 | ▼-0.307 |
| 8 | ▲0.724 | ▲0.056 | ▲0.652 | ▼-0.175 | ▲1.318 | ▲0.338 | ▼-0.078 | ▲0.471 | ▲0.418 | ▼-0.157 |
| 9 | ▲0.437 | ▼-0.197 | ▲0.574 | ▼-0.255 | ▼-0.108 | ▼-0.980 | ▼-1.157 | ▼-0.430 | ▲0.431 | ▼-0.160 |

**Fold 5**

*Train*

| 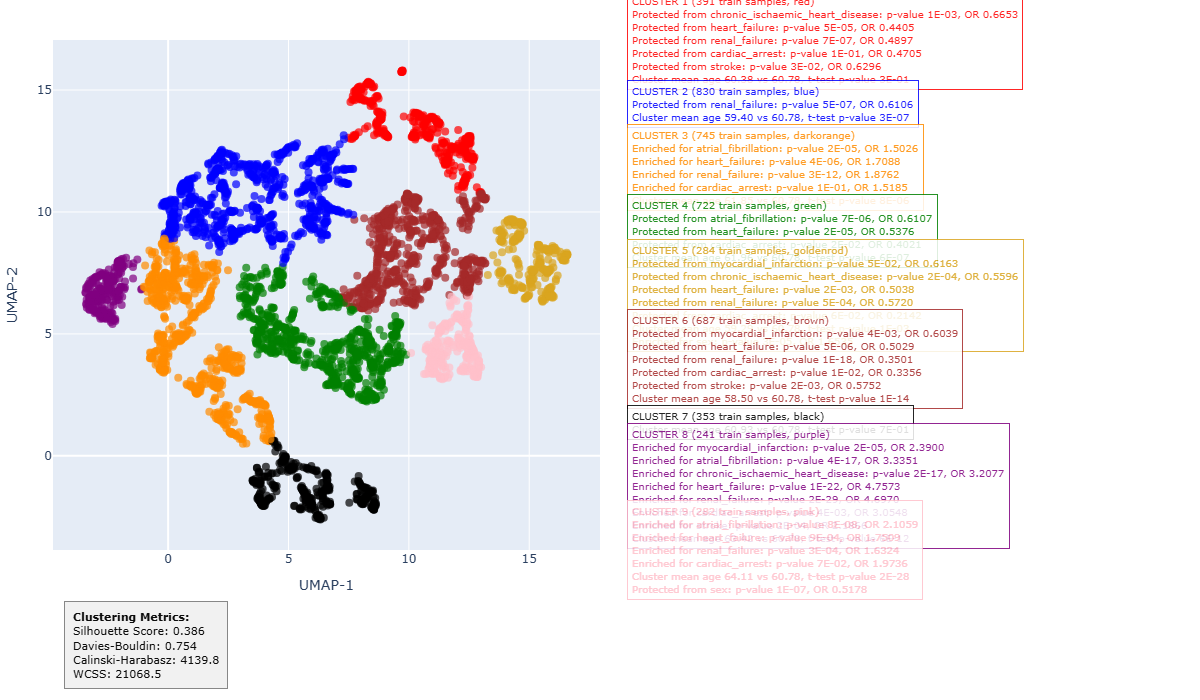 | **CLUSTER 1 (391 train samples, red)**  Protected from heart failure: p-value 5E-05, OR 0.4405 Protected from renal failure: p-value 7E-07, OR 0.4897 |
| --- | --- |
|  | **CLUSTER 2 (830 train samples, blue)** Protected from renal failure: p-value 5E-07, OR 0.6106 Cluster mean age 59.40 vs 60.78, t-test p-value 3E-07 |
|  | **CLUSTER 3 (745 train samples, dark orange)** Enriched for atrial fibrillation: p-value 2E-05, OR 1.5026 Enriched for heart failure: p-value 4E-06, OR 1.7088 Enriched for renal failure: p-value 3E-12, OR 1.8762 Cluster mean age 61.85 vs 60.78, t-test p-value 8E-06 |
|  | **CLUSTER 4 (722 train samples, green)** Protected from atrial fibrillation: p-value 7E-06, OR 0.6107 Protected from heart failure: p-value 2E-05, OR 0.5376 Protected from cardiac arrest: p-value 2E-02, OR 0.4021  Protected from cardiomyopathy: p-value 2E-3, OR 0.1798 Cluster mean age 61.96 vs 60.78, t-test p-value 6E-07 |
|  | **CLUSTER 5 (284 train samples, goldenrod)**  Protected from CIHD: p-value 2E-04, OR 0.5596 Protected from heart failure: p-value 2E-03, OR 0.5038 Protected from renal failure: p-value 5E-04, OR 0.5720 Cluster mean age 59.29 vs 60.78, t-test p-value 1E-03 Enriched for sex: p-value 3E-05, OR 1.6535 |
|  | **CLUSTER 6 (687 train samples, brown)** Protected from myocardial infarction: p-value 4E-03, OR 0.6039 Protected from heart failure: p-value 5E-06, OR 0.5029 Protected from renal failure: p-value 1E-18, OR 0.3501 Protected from cardiac arrest: p-value 1E-02, OR 0.3356 Protected from stroke: p-value 2E-03, OR 0.5752 Cluster mean age 58.50 vs 60.78, t-test p-value 1E-14 |
|  | **CLUSTER 8 (241 train samples, purple)** Enriched for myocardial infarction: p-value 2E-05, OR 2.3900 Enriched for atrial fibrillation: p-value 4E-17, OR 3.3351 Enriched for CIHD: p-value 2E-17, OR 3.2077 Enriched for heart failure: p-value 1E-22, OR 4.7573 Enriched for renal failure: p-value 2E-29, OR 4.6970 Enriched for cardiac arrest: p-value 4E-03, OR 3.0548 Enriched for stroke: p-value 2E-04, OR 2.1866 Cluster mean age 63.42 vs 60.78, t-test p-value 6E-12 |
|  | **CLUSTER 9 (282 train samples, pink)** Enriched for atrial fibrillation: p-value 8E-08, OR 2.1059 Enriched for heart failure: p-value 9E-04, OR 1.7509 Enriched for renal failure: p-value 3E-04, OR 1.6324 Cluster mean age 64.11 vs 60.78, t-test p-value 2E-28 Protected from sex: p-value 1E-07, OR 0.5178 |

**Figure S15.** Enrichments for complications, age and sex among hypertension cases from training dataset, fold 5. The silhouette score of the clustering is 0.39.

**Table S28.** Wald test *p*-values and odds ratios for the logistic regression coefficient of each cluster as a predictor of each complication reported in Figure S15 on its own and in the presence of age and sex. The *p*-values in the label, sex, and age model marked bold are significant at the *p* < 0.05 level after Benjamini-Hochberg FDR correction.

| **Cluster** | **Complication** | ***p*-value**  **(label only)** | **Odds ratio**  **(label only)** | ***p*-value**  **(label+sex+age)** | **Odds ratio**  **(label+sex+age)** |
| --- | --- | --- | --- | --- | --- |
| **1** | **heart failure** | **2E-04** | **0.44** | **5E-04** | **0.45** |
| **1** | **renal failure** | **1E-05** | **0.52** | **3E-05** | **0.54** |
| **2** | **renal failure** | **2E-07** | **0.61** | **9E-06** | **0.65** |
| **3** | **atrial fibrillation** | **1E-06** | **1.51** | **1E-04** | **1.4** |
| **3** | **heart failure** | **2E-07** | **1.7** | **4E-06** | **1.6** |
| **3** | **renal failure** | **4E-16** | **1.89** | **4E-13** | **1.8** |
| **4** | **atrial fibrillation** | **9E-06** | **0.61** | **7E-08** | **0.55** |
| **4** | **heart failure** | **5E-05** | **0.55** | **5E-06** | **0.5** |
| 4 | cardiac arrest | 7E-02 | 0.47 | 5E-02 | 0.45 |
| **4** | **cardiomyopathy** | **3E-02** | **0.3** | **3E-02** | **0.31** |
| **5** | **chronic ischaemic heart disease** | **6E-04** | **0.57** | **4E-04** | **0.55** |
| **5** | **heart failure** | **9E-03** | **0.53** | **1E-02** | **0.54** |
| **5** | **renal failure** | **2E-03** | **0.59** | **4E-03** | **0.61** |
| **6** | **myocardial infarction** | **6E-03** | **0.61** | **3E-02** | **0.68** |
| **6** | **heart failure** | **3E-05** | **0.52** | **1E-03** | **0.61** |
| **6** | **renal failure** | **7E-16** | **0.36** | **2E-12** | **0.41** |
| 6 | cardiac arrest | 5E-02 | 0.41 | 9E-02 | 0.46 |
| **6** | **stroke** | **4E-03** | **0.58** | **3E-02** | **0.66** |
| **8** | **myocardial infarction** | **4E-06** | **2.35** | **8E-05** | **2.09** |
| **8** | **atrial fibrillation** | **0E+00** | **3.28** | **1E-13** | **2.72** |
| **8** | **chronic ischaemic heart disease** | **0E+00** | **3.17** | **1E-14** | **2.79** |
| **8** | **heart failure** | **0E+00** | **4.69** | **0E+00** | **4.09** |
| **8** | **renal failure** | **0E+00** | **4.53** | **0E+00** | **4.01** |
| **8** | **cardiac arrest** | **3E-03** | **2.73** | **1E-02** | **2.4** |
| **8** | **stroke** | **2E-04** | **2.07** | **3E-03** | **1.8** |
| **9** | **atrial fibrillation** | **9E-09** | **2.09** | **5E-06** | **1.82** |
| **9** | **heart failure** | **6E-04** | **1.73** | **5E-03** | **1.57** |
| **9** | **renal failure** | **3E-04** | **1.6** | **1E-02** | **1.4** |

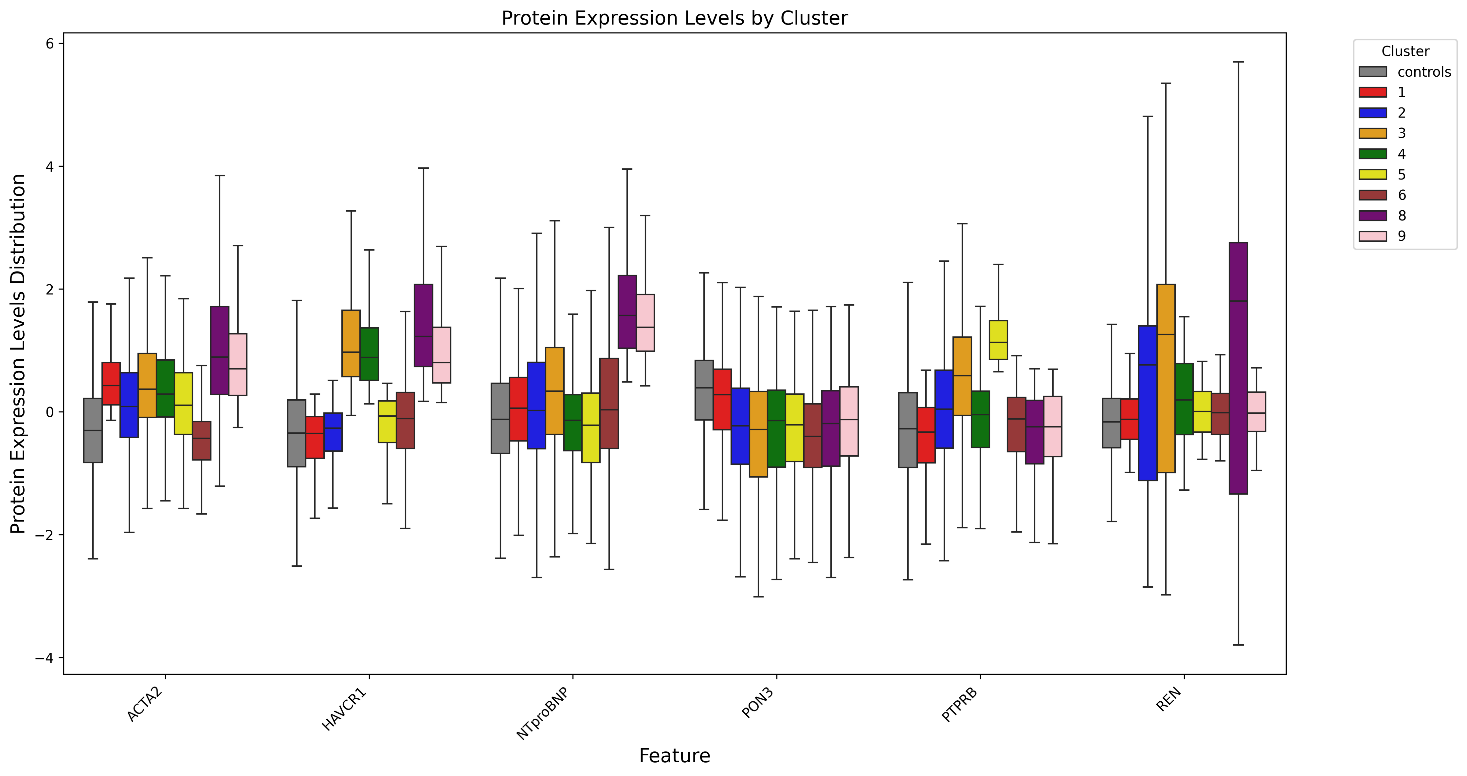
**Figure S16.** Differential expression of separating proteins among hypertension cases.

**Table S29.** Cohen's d effect sizes from Mann-Whitney U tests comparing protein expression levels between cases in each cluster and the cohort of all cases or controls. Upward-pointing arrows in red indicate up-regulated protein expression; downward-pointing arrows in blue indicate down-regulated protein expression.

| Cluster | ACTA2 | | HAVCR1 | | NTproBNP | | PON3 | | PTPRB | | REN | |
| --- | --- | --- | --- | --- | --- | --- | --- | --- | --- | --- | --- | --- |
|  | vs_controls | vs_others | vs_controls | vs_others | vs_controls | vs_others | vs_controls | vs_others | vs_controls | vs_others | vs_controls | vs_others |
| 1 | ▲0.853 | ▲0.289 | ▼-0.181 | ▼-1.044 | ▲0.184 | ▼-0.202 | ▼-0.172 | ▲0.465 | ▼-0.158 | ▼-0.719 | ▲0.066 | ▼-0.349 |
| 2 | ▲0.392 | ▼-0.208 | ▼-0.063 | ▼-1.093 | ▲0.258 | ▼-0.153 | ▼-0.673 | ▼-0.009 | ▲0.351 | ▼-0.190 | ▲0.535 | ▼-0.006 |
| 3 | ▲0.748 | ▲0.204 | ▲1.801 | ▲0.919 | ▲0.565 | ▲0.147 | ▼-0.776 | ▼-0.093 | ▲0.887 | ▲0.451 | ▲1.196 | ▲0.551 |
| 4 | ▲0.679 | ▲0.125 | ▲1.593 | ▲0.645 | ▼-0.162 | ▼-0.543 | ▼-0.685 | ▼-0.013 | ▲0.137 | ▼-0.430 | ▲0.510 | ▼-0.084 |
| 5 | ▲0.462 | ▼-0.106 | ▲0.164 | ▼-0.689 | ▼-0.140 | ▼-0.464 | ▼-0.728 | ▼-0.030 | ▲1.616 | ▲1.194 | ▲0.234 | ▼-0.242 |
| 6 | ▼-0.172 | ▼-0.880 | ▲0.237 | ▼-0.707 | ▲0.260 | ▼-0.146 | ▼-0.799 | ▼-0.102 | ▲0.021 | ▼-0.568 | ▲0.203 | ▼-0.293 |
| 7 | ▲0.685 | ▲0.123 | ▲1.866 | ▲0.848 | ▲0.214 | ▼-0.173 | ▼-0.862 | ▼-0.151 | ▲1.819 | ▲1.502 | ▲0.289 | ▼-0.214 |
| 8 | ▲1.335 | ▲0.791 | ▲2.121 | ▲1.084 | ▲2.158 | ▲1.496 | ▼-0.681 | ▲0.010 | ▼-0.040 | ▼-0.562 | ▲1.863 | ▲0.864 |
| 9 | ▲1.173 | ▲0.619 | ▲1.577 | ▲0.568 | ▲1.904 | ▲1.264 | ▼-0.577 | ▲0.101 | ▼-0.066 | ▼-0.595 | ▲0.193 | ▼-0.267 |

*Test*

| 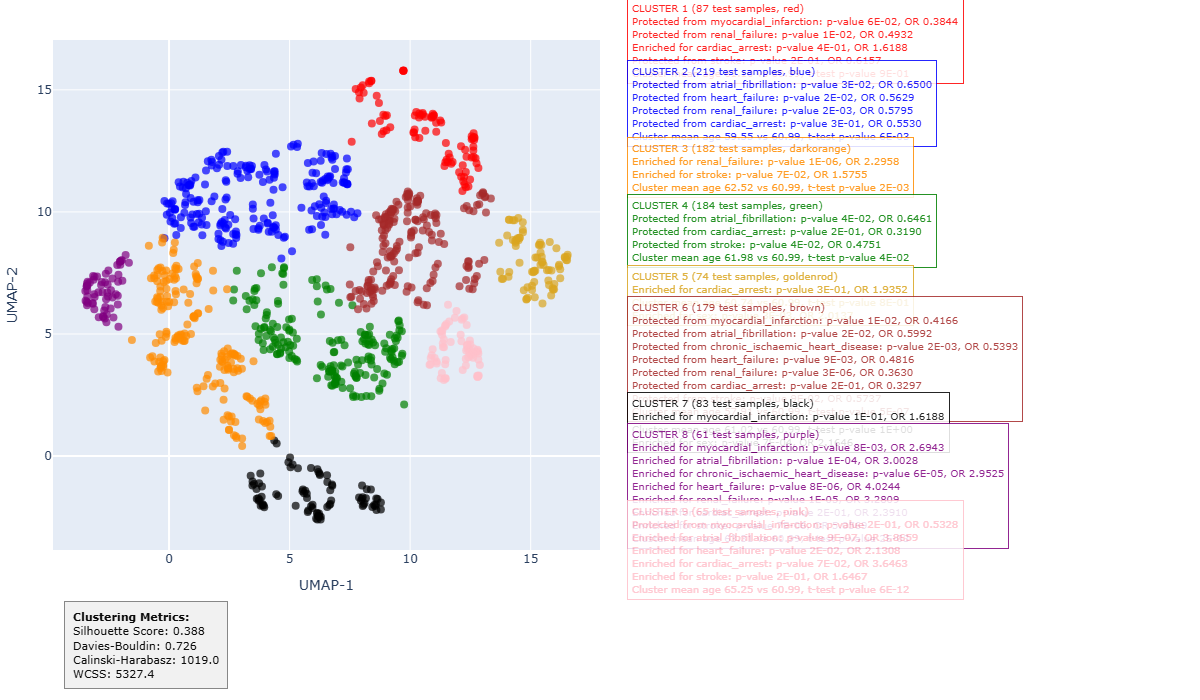 |  |  |
| --- | --- | --- |
|  |  | **CLUSTER 2 (219 test samples, blue)**  Protected from renal failure: p-value 2E-03, OR 0.5795 Cluster mean age 59.55 vs 60.99, t-test p-value 6E-03 |
|  |  | **CLUSTER 3 (182 test samples, dark orange)** Enriched for renal failure: p-value 1E-06, OR 2.2958 Cluster mean age 62.52 vs 60.99, t-test p-value 2E-03 |
|  |  | **CLUSTER 6 (179 test samples, brown)** Protected from renal failure: p-value 3E-06, OR 0.3630 Cluster mean age 57.91 vs 60.99, t-test p-value 5E-07 |
|  |  | **CLUSTER 8 (61 test samples, purple)** Enriched for myocardial infarction: p-value 8E-03, OR 2.6943 Enriched for atrial fibrillation: p-value 1E-04, OR 3.0028 Enriched for CIHD: p-value 6E-05, OR 2.9525 Enriched for heart failure: p-value 8E-06, OR 4.0244 Enriched for renal failure: p-value 1E-05, OR 3.2809 Enriched for stroke: p-value 7E-06, OR 5.0569 Cluster mean age 63.31 vs 60.99, t-test p-value 2E-03 |
|  |  | **CLUSTER 9 (65 test samples, pink)** Enriched for atrial fibrillation: p-value 9E-07, OR 3.8659 Cluster mean age 65.25 vs 60.99, t-test p-value 6E-12 |

**Figure S17.** Enrichments for complications, age and sex among hypertension cases from test dataset, fold 5. The silhouette score of the clustering is 0.39. Only complications that were statistically significant in train data according to the criteria in Enrichment analysis, and significant at the *p* < 0.05 level after Benjamini-Hochberg FDR correction are reported.

**Table S30.** Wald test *p*-values and odds ratios for the logistic regression coefficient of each cluster as a predictor of each complication reported in Figure S17 on its own and in the presence of age and sex. The *p*-values in the label, sex, and age model marked bold are significant at the *p* < 0.05 level after Benjamini-Hochberg FDR correction.

| **Cluster** | **Complication** | ***p*-value**  **(label only)** | **Odds ratio**  **(label only)** | ***p*-value**  **(label + sex + age)** | **Odds ratio**  **(label + sex + age)** |
| --- | --- | --- | --- | --- | --- |
| **2** | **renal failure** | **3E-03** | **0.59** | **1E-02** | **0.65** |
| **3** | **renal failure** | **3E-07** | **2.18** | **5E-06** | **2.02** |
| **6** | **renal failure** | **2E-05** | **0.38** | **4E-04** | **0.45** |
| **8** | **myocardial infarction** | **1E-02** | **2.37** | **1E-02** | **2.43** |
| **8** | **atrial fibrillation** | **5E-05** | **2.91** | **3E-04** | **2.63** |
| **8** | **chronic ischaemic heart disease** | **1E-04** | **2.68** | **1E-04** | **2.70** |
| **8** | **heart failure** | **5E-06** | **3.53** | **1E-05** | **3.36** |
| **8** | **renal failure** | **6E-06** | **3.15** | **4E-05** | **2.86** |
| **8** | **stroke** | **3E-06** | **4.22** | **3E-06** | **4.19** |
| **9** | **atrial fibrillation** | **5E-07** | **3.54** | **4E-05** | **2.88** |

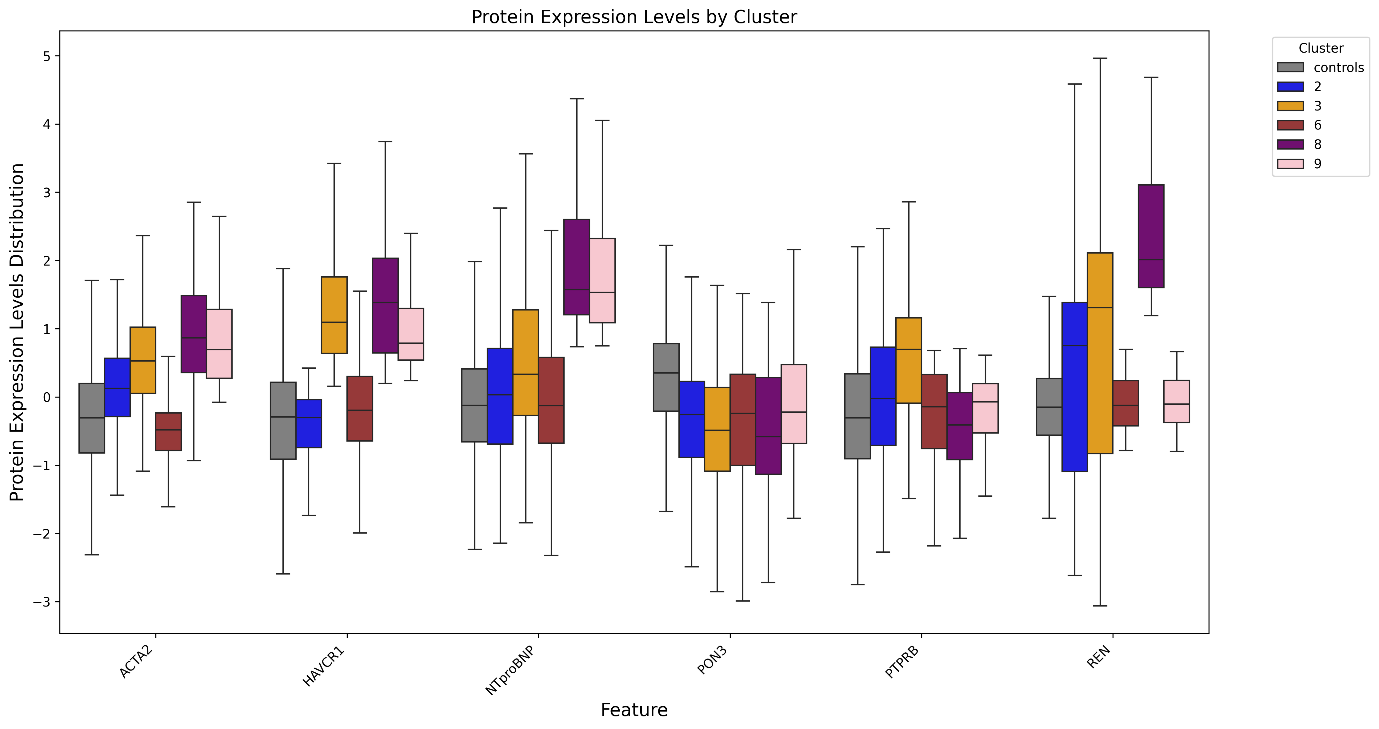

**Figure S18.** Differential expression of separating proteins among hypertension cases.

**Table S31.** Cohen's d effect sizes from Mann-Whitney U tests comparing protein expression levels between cases in each cluster and the cohort of all cases or controls. Upward-pointing arrows in red indicate up-regulated protein expression; downward-pointing arrows in blue indicate down-regulated protein expression.

| Cluster | ACTA2 | | HAVCR1 | | NTproBNP | | PON3 | | PTPRB | | REN | |
| --- | --- | --- | --- | --- | --- | --- | --- | --- | --- | --- | --- | --- |
|  | vs_controls | vs_others | vs_controls | vs_others | vs_controls | vs_others | vs_controls | vs_others | vs_controls | vs_others | vs_controls | vs_others |
| 1 | ▲0.824 | ▲0.226 | ▼-0.148 | ▼-0.977 | ▲0.279 | ▼-0.141 | ▼-0.331 | ▲0.318 | ▼-0.135 | ▼-0.658 | ▲0.030 | ▼-0.356 |
| 2 | ▲0.400 | ▼-0.252 | ▼-0.114 | ▼-1.151 | ▲0.181 | ▼-0.252 | ▼-0.643 | ▲0.034 | ▲0.337 | ▼-0.199 | ▲0.415 | ▼-0.092 |
| 3 | ▲0.931 | ▲0.405 | ▲1.865 | ▲1.027 | ▲0.726 | ▲0.264 | ▼-0.878 | ▼-0.229 | ▲0.859 | ▲0.393 | ▲1.384 | ▲0.691 |
| 4 | ▲0.820 | ▲0.262 | ▲1.606 | ▲0.700 | ▼-0.039 | ▼-0.439 | ▼-0.620 | ▲0.048 | ▲0.158 | ▼-0.400 | ▲0.270 | ▼-0.233 |
| 5 | ▲0.542 | ▼-0.067 | ▲0.161 | ▼-0.673 | ▼-0.164 | ▼-0.490 | ▼-0.606 | ▲0.067 | ▲1.713 | ▲1.258 | ▲0.260 | ▼-0.216 |
| 6 | ▼-0.232 | ▼-1.051 | ▲0.152 | ▼-0.779 | ▲0.025 | ▼-0.380 | ▼-0.747 | ▼-0.087 | ▼-0.015 | ▼-0.598 | ▲0.047 | ▼-0.383 |
| 7 | ▲0.625 | ▲0.023 | ▲1.901 | ▲0.910 | ▲0.341 | ▼-0.091 | ▼-0.704 | ▼-0.024 | ▲1.905 | ▲1.537 | ▲0.353 | ▼-0.164 |
| 8 | ▲1.349 | ▲0.802 | ▲2.026 | ▲1.018 | ▲2.377 | ▲1.592 | ▼-0.818 | ▼-0.124 | ▼-0.119 | ▼-0.623 | ▲2.623 | ▲1.357 |
| 9 | ▲1.147 | ▲0.568 | ▲1.494 | ▲0.516 | ▲2.248 | ▲1.482 | ▼-0.398 | ▲0.253 | ▲0.044 | ▼-0.464 | ▲0.097 | ▼-0.311 |
